# Predictors of long-term recreational exercise participation in adolescents and young adults

**DOI:** 10.1101/2023.04.05.23288206

**Authors:** Julie A. Morgan, Jana M. Bednarz, Ronnie Semo, Scott R. Clark, K. Oliver Schubert

**Author notes:** These authors contributed equally to this work and should be considered joint first authors (Julie A. Morgan, Jana M. Bednarz). These authors contributed equally to this work and should be considered joint senior authors (Scott R. Clark, K. Oliver Schubert).

## Abstract

The individual and societal factors influencing long-term recreational exercise participation during the transition from adolescence to young adulthood are not well explored. We modelled latent longitudinal recreational exercise trajectories spanning 8 years from age 16 to 24, and examined demographic, socioeconomic, behavioural, academic, and psychological predictors at age 15 of trajectory-group membership. We also explored whether trajectories were associated with health, mental health, and educational achievement at age 25. Finite mixture modelling was conducted with population-based longitudinal cohort study data collected from 2006-2017 by the Longitudinal Survey of Australian Youth (LSAY). The study sample comprised 9,353 students (49% female) from 356 Australian schools. Self-reported recreational exercise frequency data were collected in 2007, 2008, 2009, 2011, and 2014. Longitudinal latent trajectories of reported recreational exercise participation were estimated using group-based trajectory modelling for two scenarios: daily/guideline-adherent exercise versus non-daily exercise (model 1) and exercise at least once weekly versus exercise less than once weekly (model 2). Four distinct classes of long-term recreational exercise participation were identified for each model. Model 1: guideline-adherent exercisers (17.9% of the sample), never guideline exercisers (27.5%), guideline drop-outs (15.2%) and towards guideline (39.4%). Model 2: regular weekly (69.5% of the sample), decreasing (17.4%), increasing (4.8%), and infrequent (8.3%). In both models, predictors at age 15 for lower long-term exercise participation included female gender, lower self-efficacy, sport participation and parental socioeconomic status, and higher screen-time and academic literacy. At age 25, people in the guideline-adherent exerciser trajectory (model 1) reported better general health, whereas people in the regular weekly trajectory (model 2) had better general health and reduced rates of psychological distress, were happier with life and were more optimistic for the future relative to participants from other trajectory groups. Interventions and health-promotion activities to support sustained engagement in recreational exercise should particularly address the needs of females, people with low self-efficacy, reluctant exercisers, higher academic achievers, and youth experiencing socioeconomic disadvantage.

## 1. Introduction

The transition from adolescence to young adulthood is a critical time for the establishment of behaviours that influence health and wellbeing across the life span. Many chronic mental and physical health conditions manifest for the first time during this stage, including depression, premature diabetes, and cardiovascular disease [1-3]. Promoting positive health behaviours in young people can have benefits during youth, reduce the burden of disease in adulthood, and indirectly improve health in the next generation of children [4]. While population-level interventions for young people may powerfully impact social and economic outcomes [5], data about the development of health behaviours in 15 to 25-year-olds remain relatively sparse, prompting calls for global initiatives to improve behavioural health research in this age group [4, 6-8].

Physical exercise can confer long-term physical-and mental health benefits. In cross-sectional studies of adult populations, positive associations between exercise and cardiometabolic health, mental health, and quality of life have been reported [9, 10]. Similarly, in young people, cross-sectional studies have shown that physical exercise is associated with reduced risk for psychological distress [11], and with higher levels of bone health [12], self-esteem [11], flourishing [13], and health-related quality of life [14]. While some studies have demonstrated beneficial effects of cross-sectionally assessed exercise in youth on later adult outcomes [15, 16], distinct exercise *trajectories* during the transition from adolescence to young adulthood have received little attention [17]. Group-based trajectory modelling (GBTM) allows exploration of heterogeneity in behavioural trends over time by identifying distinct subgroups within a population. Predictors of latent group membership can be examined, and latent group membership itself may be used in turn to explain differences in particular outcomes. By identifying predictive factors for particular trajectory groups in the population, these approaches have the potential to inform health-and educational policies and aid in the targeting of preventive measures in high risk populations [18].

In the current study we applied GBTM to a large population sample to describe distinct patterns of recreational exercise undertaken outside of school or work, from the ages of 16 to 24. We then explored whether demographic, socioeconomic, behavioural, academic, and psychological factors measured at age 15 were predictive of the latent exercise trajectories describing longitudinal recreational exercise patterns, and whether trajectory group membership were associated with health, mental health, and educational achievement reported at age 25.

## 2. Materials and methods

### 2.1 Data source and participants

The Longitudinal Survey of Australian Youth (LSAY) [19] is managed by the Australian Council for Educational Research (ACER) and the Commonwealth Department of Education, Science and Training (DEST). The survey is designed to provide policy-relevant information about youth transition from school to work. Data were collected annually from 2006 at age 15 until participants turn 24 and were deposited within the Australian Data Archive (ADA) at the Australian National University. The study is jointly managed by the Australian Government Department of Education, Skills and Employment and the National Centre for Vocational Education Research (NCVER) and adheres to informed consent guidelines set by the National Health and Medical Research Council’s National Statement on the Ethical Conduct of Human Research, including written parental consent at time of enrolment of minors. Ethics approval for the LSAY survey was granted by the Australian Institute of Family Studies Ethics Committee [20]. The LSAY data are publicly available after an online registration and application process (www.ada.edu.au/). Approval was granted by The Australian Data Archive for controlled access to the LSAY 2015 cohort data (Version 1.0), and subsequently deidentified, non-reidentifiable data were available for analysis.

Data were selected from the 2006 LSAY cohort which recruited 14,170 students, at the age of 15, from 356 Australian schools involved in the Program for International Student Assessment (PISA). Participants were first assessed in 2006, followed up in 2007 by telephone, and then interviewed annually by computer assisted telephone interviews. From 2012, participants responded via an online portal until the completion of the survey in 2016.

Recreational exercise engagement was self-reported in 2007, 2008, 2009, 2011, and 2014 [19]. Longitudinal modelling was based on 9,535 participants with exercise data available. Health, mental health, and vocational outcomes were recorded in wave 10 in 2016, when participants were aged between 25 and 26 years [19].

We specified all exposure-, predictor-, and outcome variables *a priori,* based on literature review. Recreational exercise engagement was measured on a 7-point Likert-type scale, in response to the survey question “Outside study or work, how often do you play sport or do regular exercise?” (1 = every day, 2 = at least once a week (but not every day), 3 = at least once a month but less than once a week, 4 = at least once every 3 months but less often than once a month, 5 = at least once a year but less often than once every 3 months, 6 = less often than once a year, 7 = Never)[19]. Given the intervals between response categories were unequal, we derived two binary variables describing differential exercise participation, which were used in subsequent group-based trajectory modelling. The first definition defined exercise participation according to adherence to international guidelines [21], with categories of daily exercise (‘guideline-adherent’ exercise) and non-daily exercise (all other response categories; ‘non-guideline’), while the second definition differentiated between low levels of exercise participation (any response describing less than once-weekly participation; ‘infrequent’ exercise) and more regular engagement (including response categories of every day and at least once a week; ‘regular weekly’ exercise).

For both definitions, variables assessed for their association with latent trajectory groups were those previously found to be associated with exercise participation in adolescents, and included age, gender, socio-economic status (maternal and paternal International Socio-Economic Index of Occupational Status (ISEI)), indices of self-worth and self-efficacy, ethnicity (Aboriginal or Torres Strait Islander (ATSI)), attitudes towards sport, time spent playing sport, time spent watching television, academic literacy, and students interest in school [19, 22-26]. Students’ enjoyment of school was also included [19]. Candidate predictors were assessed in 2006 at age 15/16, prior to the initial period of the exercise trajectories (see Table 1).

**Table 1.** Candidate predictor variables for trajectory group membership.

| **Risk factor** | **LSAY item** | **Year measured** | **Unit/values (range)** | **Proportion of data missing**<br>**(as % of N=9353)** |
| --- | --- | --- | --- | --- |
| Sex | Gender | 2006 | (1) Male<br>(2) Female | 0 (no missing data) |
| International Socio-Economic Index of Occupational Status (proxy for SES) | Father's ISES score | 2006 | 1-100 | 7.45% |
|  | Mother's ISES score | 2006 | 1-100 | 11.1% |
| Self-efficacy | "Compared with most students in your year level, how well are you doing in your subjects overall?" | 2006 | (1) Below average<br>(2) About average<br>(3) Above average | 0.77% |
| Self-worth | "My school is a place where I know I can do well enough to be successful" | 2006 | (1) Agree<br>(2) Disagree | 0.71% |
|  | "I am a success as a student" | 2006 | (1) Agree<br>(2) Disagree | 1.28% |
|  | “My school is a place where teachers give me marks I deserve” | 2006 | (1) Agree<br>(2) Disagree | 0.97% |
| Ethnicity | ATSI-status | 2006 | (1) Indigenous<br>(2) Non-indigenous | 0 (no missing data) |
| Sports participation | On average, how many hours per week do you spend playing sport? | 2006 | minutes per week | 2.11% |
| Screen time: TV | On average, how many hours do you spend each week on watching TV? | 2006 | minutes per week | 1.85% |
| Enjoyment of school | My school is a place where I get enjoyment from being here | 2006 | (1) Agree<br>(2) Disagree | 0.92% |
| Academic aptitude | Plausible value in maths | 2006 | 670-3950 | 0 (no missing data) |
|  | Plausible value in reading | 2006 | 670-3950 | 0 (no missing data) |
|  | Plausible value in science | 2006 | 670-3950 | 0 (no missing data) |
|  | Academic literacy (sum of maths, reading and science scores) | 2006 | 2010-11850 | 0 (no missing data) |
| Work at school | My school is a place where: I am given the chance to do work that really interests me | 2006 | (1) Agree<br>(2) Disagree | 0.74% |

Note: Maximum plausible upper limits were imposed for sports participation (responses >40 hours/week excluded, n=91) and TV time (responses > 74 hours/week excluded n=42).

Several self-reported indicators of mental health, physical health, and vocational attainment were measured in 2016 (at age 25). These included the Kessler-6 psychological distress score (K6), a validated measure widely used as an index of mental health in Australian population-based studies where comprehensive assessments are not feasible [19, 27]. Other measures taken at this timepoint included a single item from the SF-36 to qualify general health (“How would you say your health is?”) [19, 28], and items regarding on satisfaction with life and view of one’s future, completion of high school, completion of a post school qualification, and participation in the labour force [19] (see Table 2). K6 scores were dichotomised into either 0 = lower risk of mental illness (scores from 6 - 18) or 1 = greater risk of mental illness (scores > 19) [29]. Predictor variables of exercise trajectory groups were included in outcome analyses to control for potential confounding.

**Table 2.** Outcome variables assessed at age 25.

| Outcome | LSAY item | Year measured | Unit/values (range) |
| --- | --- | --- | --- |
| General health | “In general, how would you say your health is?”<br>(from SF-36) | 2016 | (1) excellent<br>(2) very good<br>(3) fair<br>(4) poor |
| Greater risk of mental illness | Kessler-6 (K6) questionnaire | 2016 | (1) lower risk of mental illness (score of 8-18)<br>(2) greater risk of mental illness (score of 19-30) |
| Life satisfaction | “How happy are you with your life as a whole?” | 2016 | (1) very happy or happy<br>(2) unhappy or very unhappy |
| View of one's future | “How happy are you with your future?” | 2016 | (1) very happy or happy<br>(2) unhappy or very unhappy |
| Completion of high school | Completed Year 12 or Certificate II or higher | 2016 | (1) Year 12 completed<br>(2) Year 12 not completed |
| Completion of post-school qualification | Completed any post-school qualification | 2016 | (1) Qualification completed<br>(2) Qualification not completed |
| Participation in the labour force | Derived variable | 2016 | (1) Participating in labour force (employed)<br>(2) Not participating in labour force (unemployed) |

### 2.2 Statistical methods

We applied group-based trajectory modelling (GBTM) to the two dichotomised exercise-frequency variables described above to explore exercise behaviour over time (*model 1*: daily exercise/*guideline* vs less than daily/*non-guideline; model 2*: at least 1x per week/*regular weekly* vs less than 1x per week/*infrequent*). GBTM is a specialised application of finite mixture modelling that identifies distinct subgroups of individuals who follow similar trajectories of a repeatedly-measured behaviour [30]. Each trajectory is described as a polynomial function of time or age. To determine the optimal number of trajectory groups suggested by the observed data, and their shape (polynomial-order), a series of models with 1 to 4 groups of varying polynomial order were implemented using the *traj* add-on for Stata [31], separately for each definition of exercise engagement. A Bernoulli distribution was assumed since the exercise behaviour measures were binary. In the absence of any *a priori* hypotheses surrounding the number and shape of trajectory groups, for each measure we considered all possible models of group size ranging from one to four, and all possible combinations of zero-order (a constant), linear and quadratic polynomials for the trajectory shapes. Models with trajectory groups containing less than 1% of the sample were excluded, and the final models were selected based on goodness of fit as assessed using the Bayesian Information Criteria (BIC). The *traj* add-on uses full information maximum likelihood to handle missing data, such that parameters will be asymptotically unbiased under an assumption that missingness is not related to the unobserved outcomes (i.e. trajectory group assignment) [32]. There was little evidence against the null hypothesis that the exercise data were Missing Completely At Random (MCAR) from Little’s test (χ_(33)_^2^=20.2, p=0.961), such that the assumption that attrition is independent of trajectory group is reasonable.

For each of the two final models, participants were then assigned to the trajectory group for which they had the highest probability for membership, and the pre-specified baseline factors were investigated for their association with trajectory group membership post-hoc using multinomial logistic regression. All predictor variables had low amounts of missing data (<5%).

Associations between the identified trajectory groups in each of the two final models and longer-term outcomes were assessed using logistic regression (categorical outcomes), and linear regression (continuous outcomes), adjusted for potential confounders (gender, indigeneity, student socio-economic status (SSES), father ISEI, self-efficacy, academic achievement, and sport participation, TV time, and enjoyment of school at baseline in 2006) [19]. Standard errors were adjusted to account for clustering of participants by school. Goodness of fit was evaluated using the Hosmer-Lemeshow test for logistic regression models, and by visual inspection of residual plots for normality and homoscedasticity for linear regression models. All analyses were conducted in Stata (version 15 StataCorp Texas) including use of the ‘traj’ add-on[31]. The level of statistical significance was set to 0.05.

## 3. Results

### 3.1 Group Based Trajectory Modelling identified four distinct patterns of recreational exercise participation over the 8-year period from the age of 16 to 24

Exercise Trajectory Model 1 evaluated the binary variable *guideline* vs *non-guideline* exercisers. Proportions of young people falling into these categories at each assessment time point are shown in S1 Fig. The final trajectory model 1, as determined by the Bayesian information criterion (BIC), was a four-group model, with groups of polynomial order 0, 0, 2 and 2, respectively (Fig 1). All trajectory subgroups included more than 1% of the sample. The two groups showing stable probabilities of exercise over time were labelled *guideline* (a high probability of daily exercise over time; 17.9% of the sample) and *never guideline* (low probability of daily exercise; 27.5% of the sample). The two groups with changes in probability over time were labelled *guideline drop-outs* (15.2% of the sample) and *towards guideline* (39.4% of the sample).

Exercise Trajectory Model 2 used the binary variable *infrequent* vs *regular weekly* exercisers. Proportions of young people falling into these categories at each assessment time point are shown in S2 Fig. As determined by the BIC, model 2 was also a four-group model, with groups of polynomial order 0, 0, 2 and 2, respectively (Fig 2). All exercise trajectory subgroups included more than 1% of the sample. We labelled the two stable groups *infrequent* exercisers (with high probability of very little exercise over time, 8.3% of the sample) and *regular weekly* exercisers (69.5% of the sample). The two groups showing change in behaviour over time were labelled *declining-exercisers* (17.4%), and *increasing-exercisers* (4.8%), respectively.

**Legend Figs 1 and 2.**
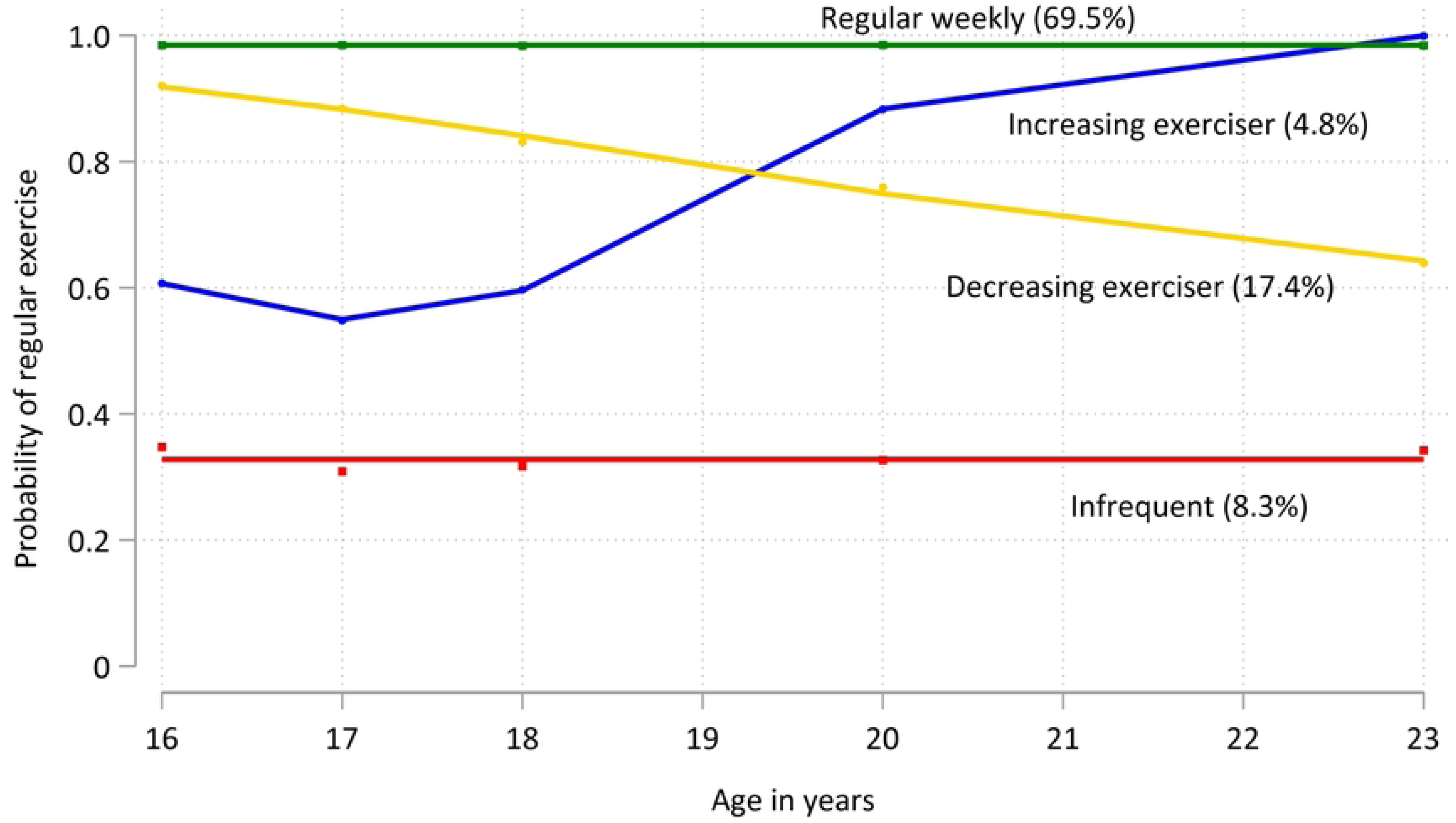
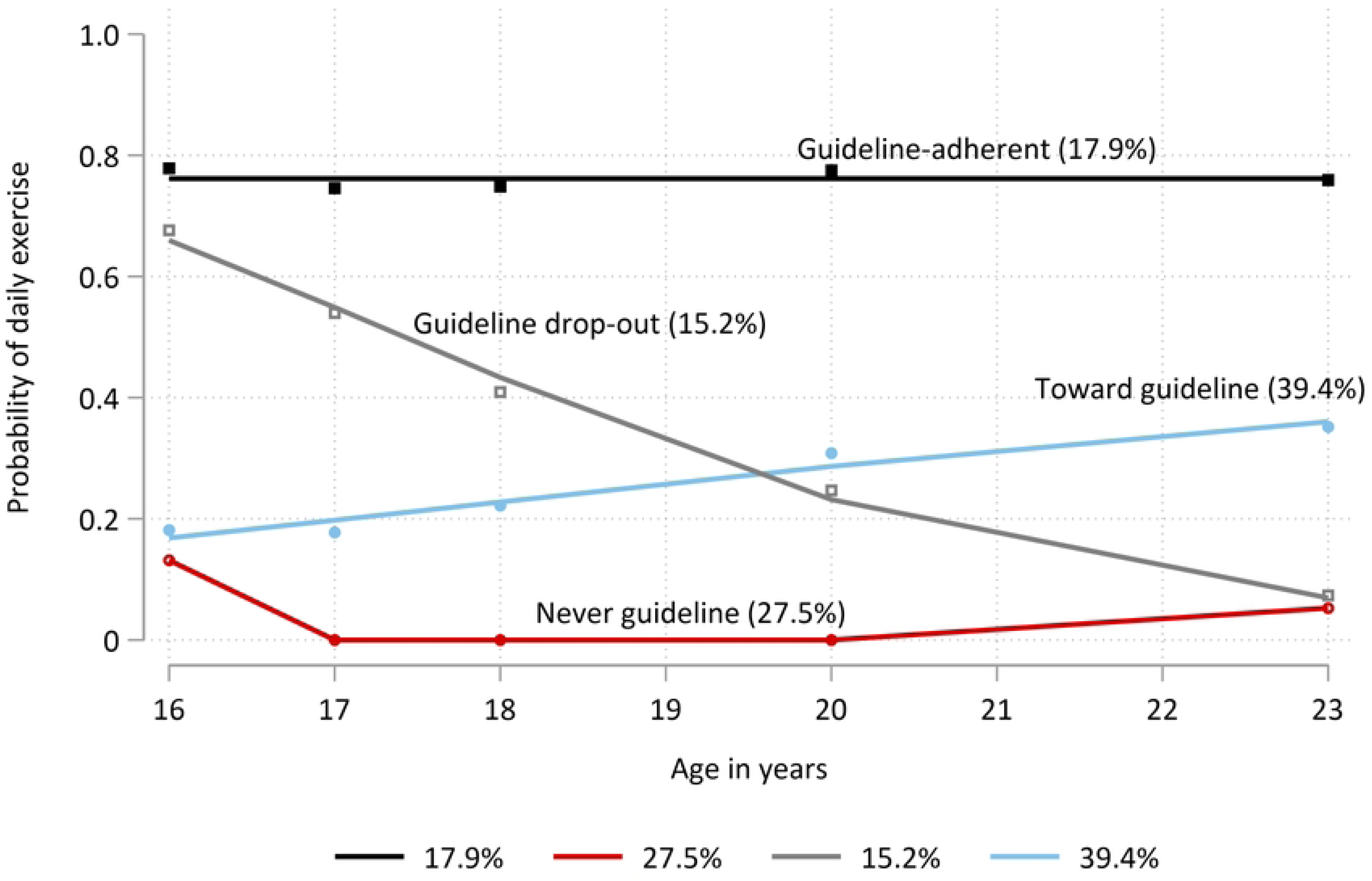
Models for 8-year physical exercise trajectories from age 16 to 24. **1) : Exercise Trajectory Model 1.** Of the 9,353 young people included in the model, 17.9 % had a high probability of consistently meeting the WHO guideline recommendation of daily exercise, over 8 years (*guideline,* black line). 27.5% were unlikely to ever meet recommendations (*never guideline*, dark red line). 15.2% of the sample were initially meeting daily exercise requirements but dropped out over time (*guideline drop-outs*, grey line). 39.4% of young people had a low initial probability of meeting guideline requirements but were more likely as time progressed (*towards guideline*, blue line). **2) Exercise Trajectory Model 2.** 8.3% of young people had a persistently low probability of meaningful recreational physical exercise engagement, reporting less than one occasion per week (*infrequent*, red line). In contrast, 69.5% had a high probability of engaging consistently in more regular exercise (*regular weekly*, green line). 17.4% of the sample had an initially high probability of regular exercise that declined over time (*decreasing exerciser*, yellow line). 4.8% had a modest initial probability of regular exercise that increased from age 17 (*increasing-exerciser*, blue line).

### 3.2 Predictors at age 15 for long-term adherence to exercise guidelines (daily exercise)

Predictors for long-term adherence to daily exercise were those factors that reduced the risk of membership in a non-guideline trajectory group (reported as the adjusted Relative Risk Ratio (aRRR) for membership to a non-guideline group vs the guideline-adherent group, 95% CI; p). More time playing sport was a strong predictor of long-term maintenance of daily exercise: aRRR for each hour playing sport per week for *never guideline* vs *guideline*=0.86, 95% CI 0.84, 0.87; p<.0001; for *guideline dropout* vs *guideline* =0.98, 95% CI 0.97, 0.99; p<0.01; for *towards guideline* vs *guideline*=0.91 95% CI 0.90, 0.92; p<0.001 (Figs 3 - 5, Table 3).

**Table 3.** Summary statistics for continuous predictors for model 1 trajectory groups.

| Characteristic | Assigned Trajectory Group |
| --- | --- |

|  | Guideline-adherent<br>(n=1948) | Never<br>guideline<br>(n=3176) | Guideline<br>drop-out<br>(n=612) | Towards<br>guideline<br>(n=3617) |
| --- | --- | --- | --- | --- |
| **International Socio-Economic Index of**<br>**Occupational Status (ISEI)** |  |  |  |  |
| Father's ISEI score (mean ± SD) | 51.7 ± 22.0 | 50.8 ± 21.5 | 52.2 ± 21.1 | 50.9 ± 22.3 |
| Mother's ISEI score (mean ± SD) | 53.5 ± 21.6 | 53.0 ± 21.8 | 54.4 ± 21.4 | 53.7 ± 22.5 |
| **Time spent each week playing sport**<br>**(hours) at baseline (mean ± SD)** | 9.0 ± 6.9 | 4.5 ± 4.2 | 8.0 ± 6.1 | 5.6 ± 5.3 |
| **Time spent each week watching TV**<br>**(hours) at baseline (mean ± SD)** | 10.3 ± 8.9 | 10.8 ± 9.2 | 10.5 ± 9.0 | 10.9 ± 9.2 |
| **Academic literacy at baseline (mean ± SD)** |  |  |  |  |
| Plausible value in maths, science and reading<br>combined | 7870 ± 1243 | 8199 ± 1220 | 8156 ±<br>1200 | 7841 ±<br>1250 |

Conversely, factors that increased risk of membership in a non-guideline exercise trajectory group (reported as aRRR for membership to a non-guideline vs guideline-adherent group, 95% CI; p) were: female gender: aRRR for *never guideline* vs *guideline* =1.76, 95% CI 1.53, 2.02; p<.0001; for *towards guideline* vs *guideline* =1.61, 95% CI 1.41, 1.84; p=0.001), lower academic self-efficacy (relative to higher self-efficacy): aRRR for *never guideline* vs *guideline* =1.20, 95% CI 1.02, 1.39; p=0.025; for *towards guideline* vs *guideline* =1.25, 95% CI 1.09, 1.43; p<0.001), greater academic literacy: aRRR per 1000 point increase for *never guideline* vs *guideline* =1.32, 95% CI 1.24, 1.40; p<0.001; *guideline dropout* vs *guideline* =1.17, 95% CI 1.06, 1.30; p=0. and more time watching TV: aRRR per 5 hour increase per week for *never guideline* vs *guideline*=1.14, 95% CI 1.09, 1.20; p<.0001; *towards guideline* vs *guideline* =1.10 95% CI 1.05, 1.15; p<0.001) (Figs 3 - 5, Tables 3 and 4).

**Legend Figs 3 - 5.**
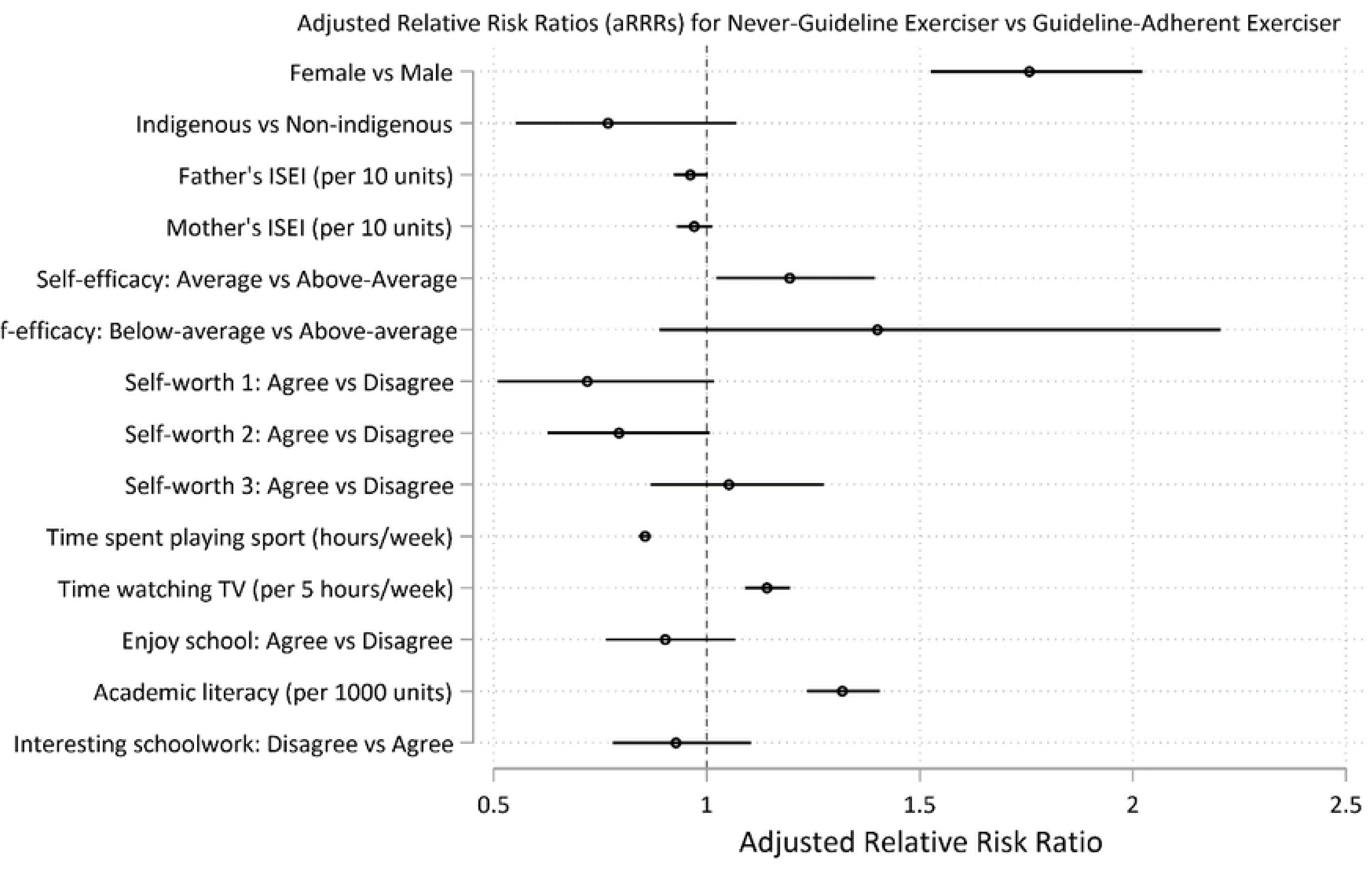
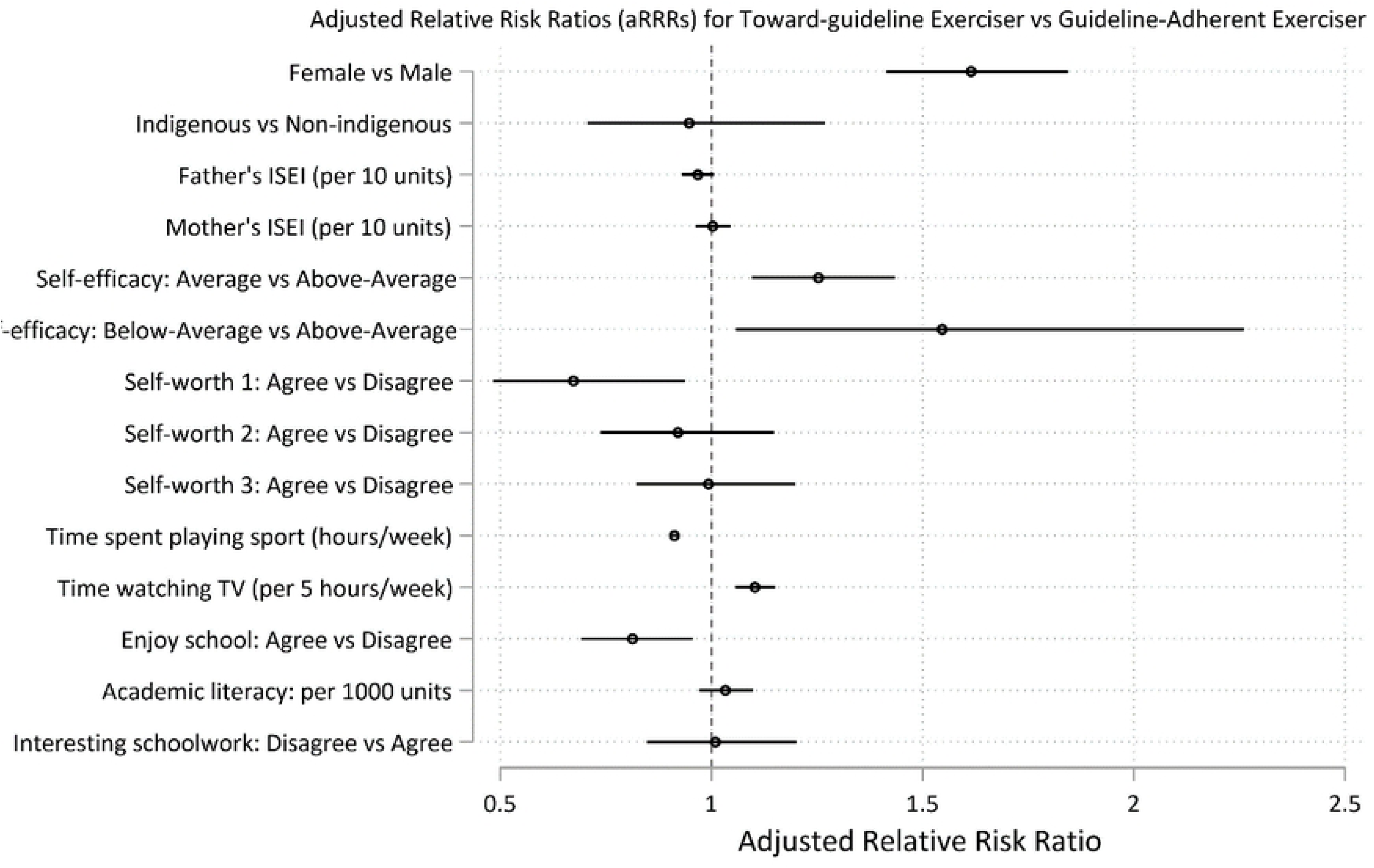
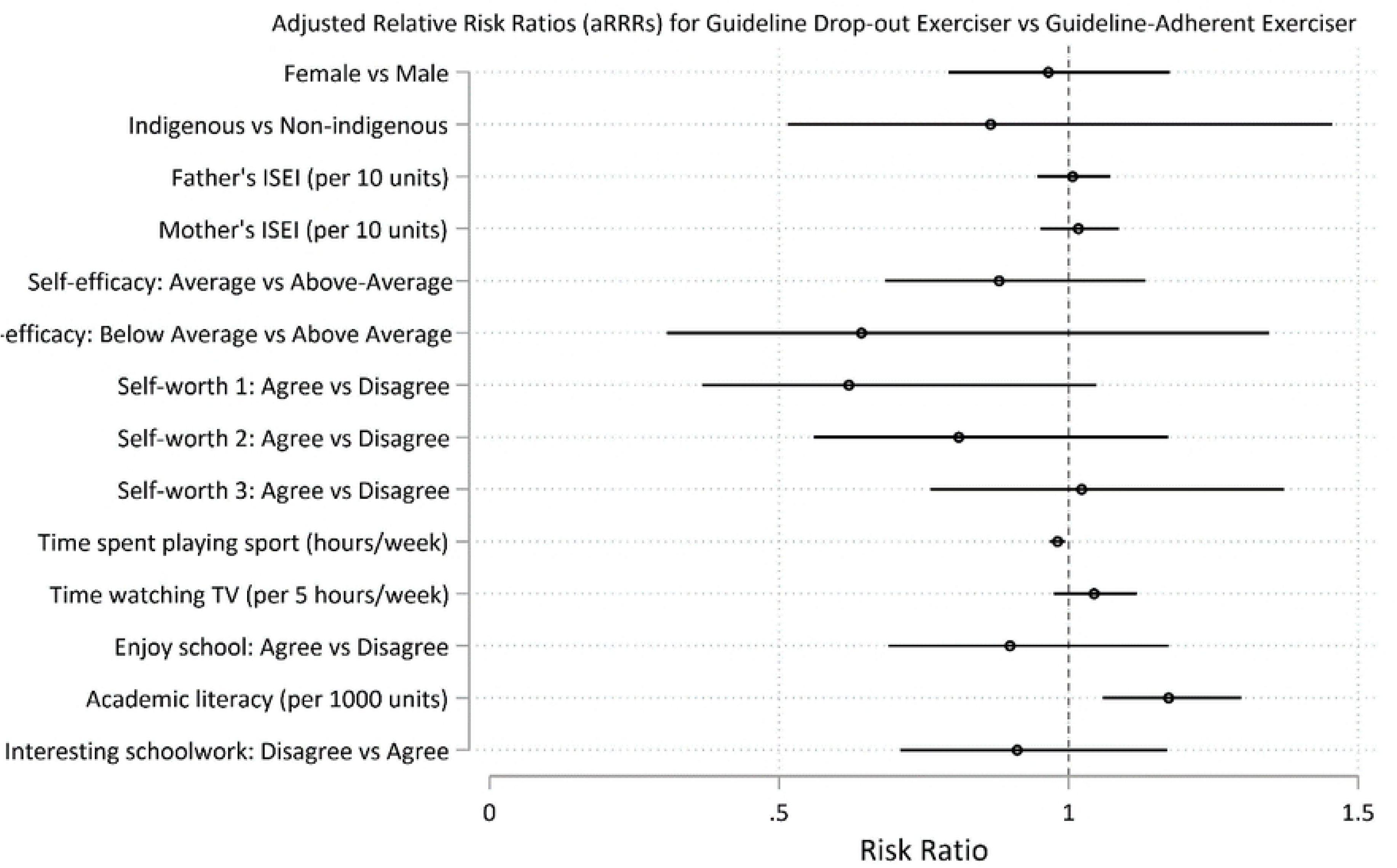
Predictors at age 15 for long-term recreational physical exercise participation between the ages of 16 and 24. Predictors for guideline-adherent (=daily) exercise. Predictors are shown as adjusted Relative Risk Ratios (aRRRs) for falling into the *never guideline* exerciser trajectory (Fig 3), the *towards guideline* trajectory (Fig 4) and the *guideline drop-out* trajectory (Fig 5), compared to the risk of following the *guideline* exercise trajectory.

**Table 4.** Summary statistics for binary and categorical predictors for model 1 trajectory groups. Percentages may not equal 100 due to rounding.

| Characteristic | Assigned Trajectory Group<sup>^</sup> |  |  |  |  |  |  |  |
| --- | --- | --- | --- | --- | --- | --- | --- | --- |
|  | Guideline-adherent<br>(n=1948) |  | Never<br>guideline<br>(n=3176) |  | Guideline<br>drop-out<br>(n=612) |  | Towards<br>guideline<br>(n=3617) |  |
|  | Freq | (%) | Freq | (%) | Freq | (%) | Freq | (%) |
| **Gender** |  |  |  |  |  |  |  |  |
| Male | 1187 | (60.9) | 1289 | (40.6) | 369 | (60.3) | 1616 | (44.7) |
| Female | 761 | (39.1) | 1887 | (59.4) | 243 | (39.7) | 2001 | (55.3) |
| **Indigenous Status** |  |  |  |  |  |  |  |  |
|  | Guideline- |  | Never |  | Guideline |  | Towards |  |
|  | adherent |  | guideline |  | drop-out |  | guideline |  |
|  | (n=1948) |  | (n=3176) |  | (n=612) |  | (n=3617) |  |
|  | Freq | (%) | Freq | (%) | Freq | (%) | Freq | (%) |
| Non-indigenous | 1824 | (93.6) | 3036 | (95.6) | 583 | (95.3) | 3392 | (93.8) |
| Indigenous | 124 | (6.4) | 140 | (4.4) | 29 | (4.7) | 225 | (6.2) |
| **Self-efficacy** |  |  |  |  |  |  |  |  |
| *“Compared with most students in your year level, how well are you doing in your subjects overall?”* |  |  |  |  |  |  |  |  |
| Below average | 71 | (3.6) | 130 | (4.1) | 17 | (2.8) | 183 | (5.1) |
| About average | 747 | (38.4) | 1300 | (40.9) | 209 | (34.2) | 1639 | (45.3) |
| Above average | 1113 | (57.1) | 1721 | (54.2) | 384 | (62.8) | 1767 | (48.9) |
| Missing/unknown | 17 | (0.9) | 25 | (0.8) | 2 | (0.33) | 28 | (0.77) |
| **Self-worth** |  |  |  |  |  |  |  |  |
| *“My school is a place where I know I can do well enough to be successful”* |  |  |  |  |  |  |  |  |
| Agree | 1834 | (94.2) | 2972 | (93.6) | 578 | (94.4) | 3324 | (91.9) |
| Disagree | 97 | (5.0) | 184 | (5.8) | 34 | (5.6) | 264 | (7.3) |
| Missing/unknown | 17 | (0.9) | 20 | (0.6) | 0 | (0) | 29 | (0.8) |
| *“I am a success as a student”* |  |  |  |  |  |  |  |  |
| Agree | 1684 | (86.5) | 2692 | (84.8) | 528 | (86.3) | 2985 | (82.5) |
| Disagree | 236 | (12.1) | 449 | (14.1) | 77 | (12.6) | 582 | (16.1) |
| Missing/unknown | 28 | (1.4) | 35 | (1.1) | 7 | (1.1) | 50 | (1.4) |
| *“My school is a place where teachers give me marks I deserve”* |  |  |  |  |  |  |  |  |
| Agree | 1592 | (81.7) | 2714 | (85.5) | 515 | (84.2) | 2941 | (81.3) |
|  | Guideline- |  | Never |  | Guideline |  | Towards |  |
|  | adherent |  | guideline |  | drop-out |  | guideline |  |
|  | (n=1948) |  | (n=3176) |  | (n=612) |  | (n=3617) |  |
|  | Freq | (%) | Freq | (%) | Freq | (%) | Freq | (%) |
| Disagree | 333 | (17.1) | 432 | (13.6) | 95 | (15.5) | 640 | (17.7) |
| Missing/unknown | 23 | (1.2) | 30 | (0.9) | 2 | (0.3) | 36 | (1.0) |
| **Enjoyment of school** |  |  |  |  |  |  |  |  |
| *“My school is a place where I get enjoyment from being here”* |  |  |  |  |  |  |  |  |
| Agree | 1471 | (75.5) | 2341 | (73.7) | 459 | (75.0) | 2549 | (70.5) |
| Disagree | 459 | (23.6) | 811 | (25.5) | 150 | (24.5) | 1027 | (28.4) |
| Missing/unknown | 18 | (0.9) | 24 | (0.8) | 3 | (0.5) | 41 | (1.1) |
| *“At school I am given the chance to do interesting work”* |  |  |  |  |  |  |  |  |
| Agree | 1453 | (74.6) | 2414 | (76.0) | 478 | (78.1) | 2607 | (72.1) |
| Disagree | 479 | (24.6) | 743 | (23.4) | 131 | (21.4) | 979 | (27.1) |
| Missing/unknown | 16 | (0.8) | 19 | (0.6) | 3 | (0.49) | 31 | (0.9) |

### 3.3 Risk factors at age 15 for insufficient long-term recreational exercise participation (less than once weekly)

At age 15, the risk factors for membership in one of the three groups engaging in less exercise (infrequent exercisers, increasing exercisers, and declining exercisers) relative to the regular weekly group were as follows: female gender (aRRR for *infrequent exercisers* =1.55, 95% CI 1.30, 1.85; p<.0001 ; aRRR for *declining exercisers* =1.27, 95% CI 1.12, 1.43; p<.0001; aRRR for *increasing exercisers* =1.47, 95% CI 1.18, 1.83; p=0.001), increased TV time (for every 5 hours of TV per week, aRRR for *infrequent exercisers* =1.13, 95% CI 1.08, 1.18; p<.0001 ; aRRR for *declining exercisers* =1.06, 95% CI 1.02, 1.09; p=0.001; aRRR for *increasing exercisers* =1.11, 95% CI 1.05, 1.17; p<.0001), and average levels of academic self-efficacy (versus above-average self-efficacy) (aRRR for *infrequent exercisers* =1.32, 95% CI 1.10, 1.18; p=0.003; aRRR for *declining exercisers* =1.18, 95% CI 1.03, 1.35; p=0.016; aRRR for *increasing exercisers* =1.27, 95% CI 1.02, 1.58; p=0.035). Higher sports participation at age 15 decreased the risk of being in the lower exercise trajectories (for each hour spent playing sport per week, aRRR for *infrequent exercisers* =0.69, 95% CI 0.65, 0.73; p<.0001 ; aRRR for *declining exercisers* =0.91, 95% CI 0.90, 0.93; p<.0001; aRRR for *increasing exercisers* =0.81, 95% CI 0.77, 0.85; p<.0001) (Figs 6 – 8, Tables 5 and 6).

**Table 5.**
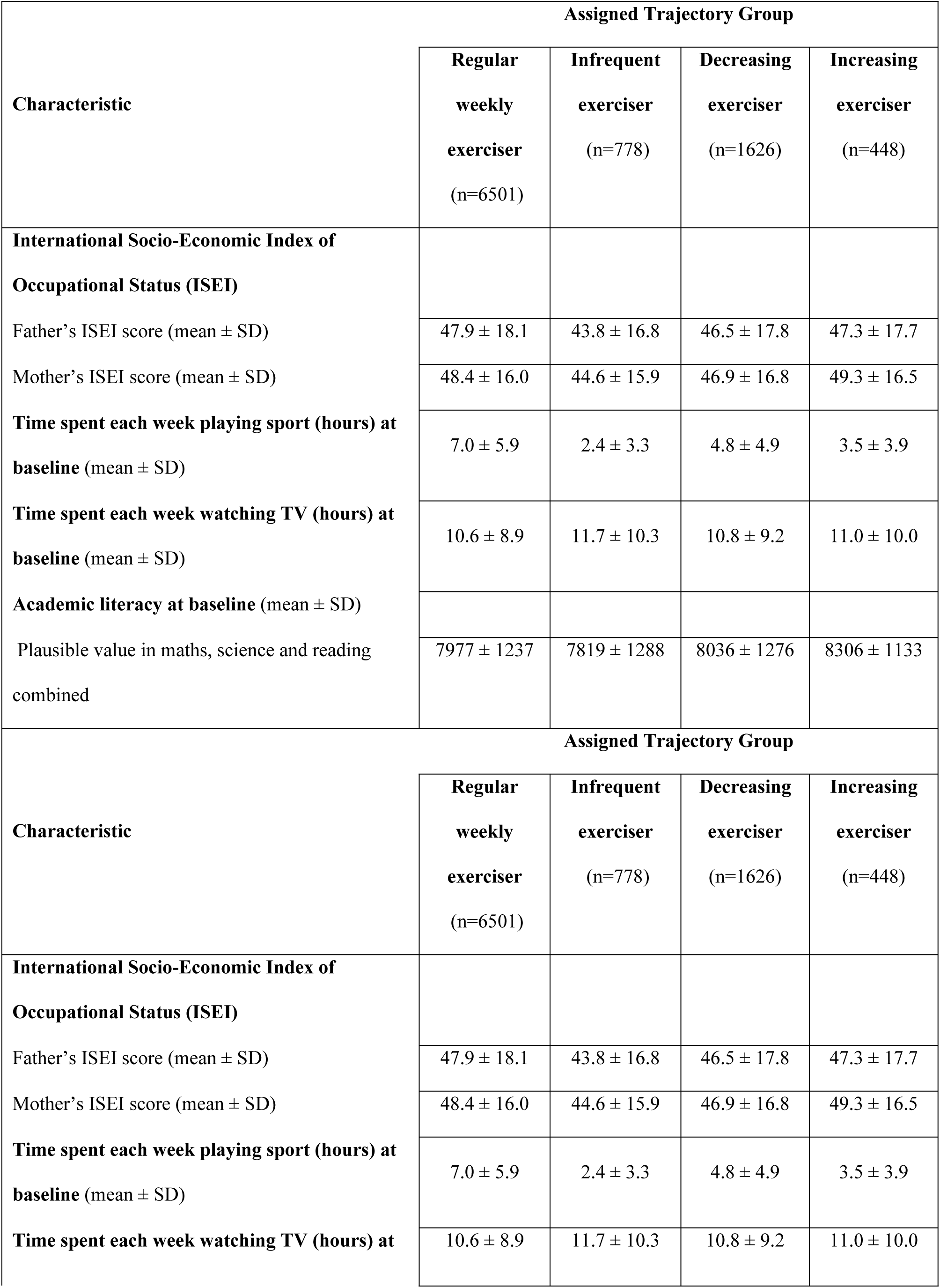
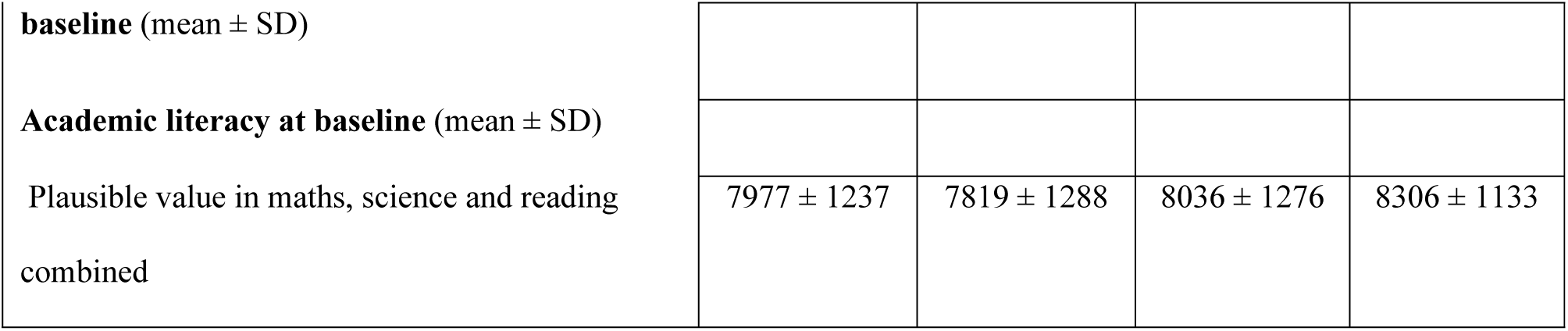
Summary statistics for continuous variables for model 2 trajectory groups.

| Characteristic | Assigned Trajectory Group |  |  |  |
| --- | --- | --- | --- | --- |
|  | Regular<br>weekly<br>exerciser<br>(n=6501) | Infrequent<br>exerciser<br>(n=778) | Decreasing<br>exerciser<br>(n=1626) | Increasing<br>exerciser<br>(n=448) |
| **International Socio-Economic Index of Occupational Status (ISEI)** |  |  |  |  |
| Father's ISEI score (mean ± SD) | 47.9 ± 18.1 | 43.8 ± 16.8 | 46.5 ± 17.8 | 47.3 ± 17.7 |
| Mother's ISEI score (mean ± SD) | 48.4 ± 16.0 | 44.6 ± 15.9 | 46.9 ± 16.8 | 49.3 ± 16.5 |
| **Time spent each week playing sport (hours) at baseline** (mean ± SD) | 7.0 ± 5.9 | 2.4 ± 3.3 | 4.8 ± 4.9 | 3.5 ± 3.9 |
| **Time spent each week watching TV (hours) at baseline** (mean ± SD) | 10.6 ± 8.9 | 11.7 ± 10.3 | 10.8 ± 9.2 | 11.0 ± 10.0 |
| **Academic literacy at baseline** (mean ± SD) | | | | |
| Plausible value in maths, science and reading combined | 7977 ± 1237 | 7819 ± 1288 | 8036 ± 1276 | 8306 ± 1133 |
| Characteristic | Assigned Trajectory Group |  |  |  |
|  | Regular<br>weekly<br>exerciser<br>(n=6501) | Infrequent<br>exerciser<br>(n=778) | Decreasing<br>exerciser<br>(n=1626) | Increasing<br>exerciser<br>(n=448) |
| **International Socio-Economic Index of Occupational Status (ISEI)** |  |  |  |  |
| Father's ISEI score (mean ± SD) | 47.9 ± 18.1 | 43.8 ± 16.8 | 46.5 ± 17.8 | 47.3 ± 17.7 |
| Mother's ISEI score (mean ± SD) | 48.4 ± 16.0 | 44.6 ± 15.9 | 46.9 ± 16.8 | 49.3 ± 16.5 |
| **Time spent each week playing sport (hours) at baseline** (mean ± SD) | 7.0 ± 5.9 | 2.4 ± 3.3 | 4.8 ± 4.9 | 3.5 ± 3.9 |
| **Time spent each week watching TV (hours) at baseline** (mean ± SD) | 10.6 ± 8.9 | 11.7 ± 10.3 | 10.8 ± 9.2 | 11.0 ± 10.0 |
| **baseline** (mean ± SD) |  |  |  |  |
| **Academic literacy at baseline** (mean ± SD) |  |  |  |  |
| Plausible value in maths, science and reading combined | 7977 ± 1237 | 7819 ± 1288 | 8036 ± 1276 | 8306 ± 1133 |

**Table 6.**
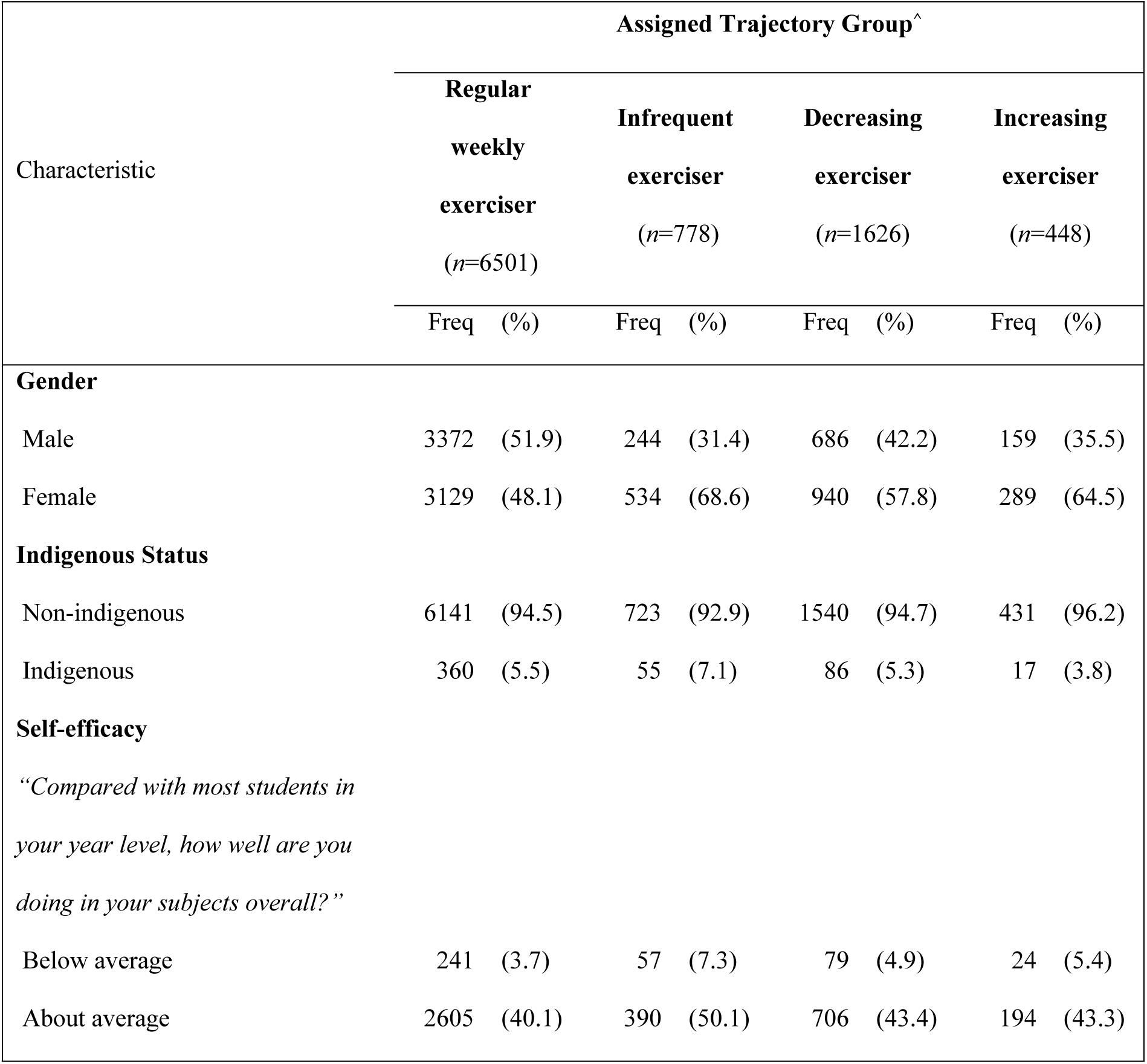
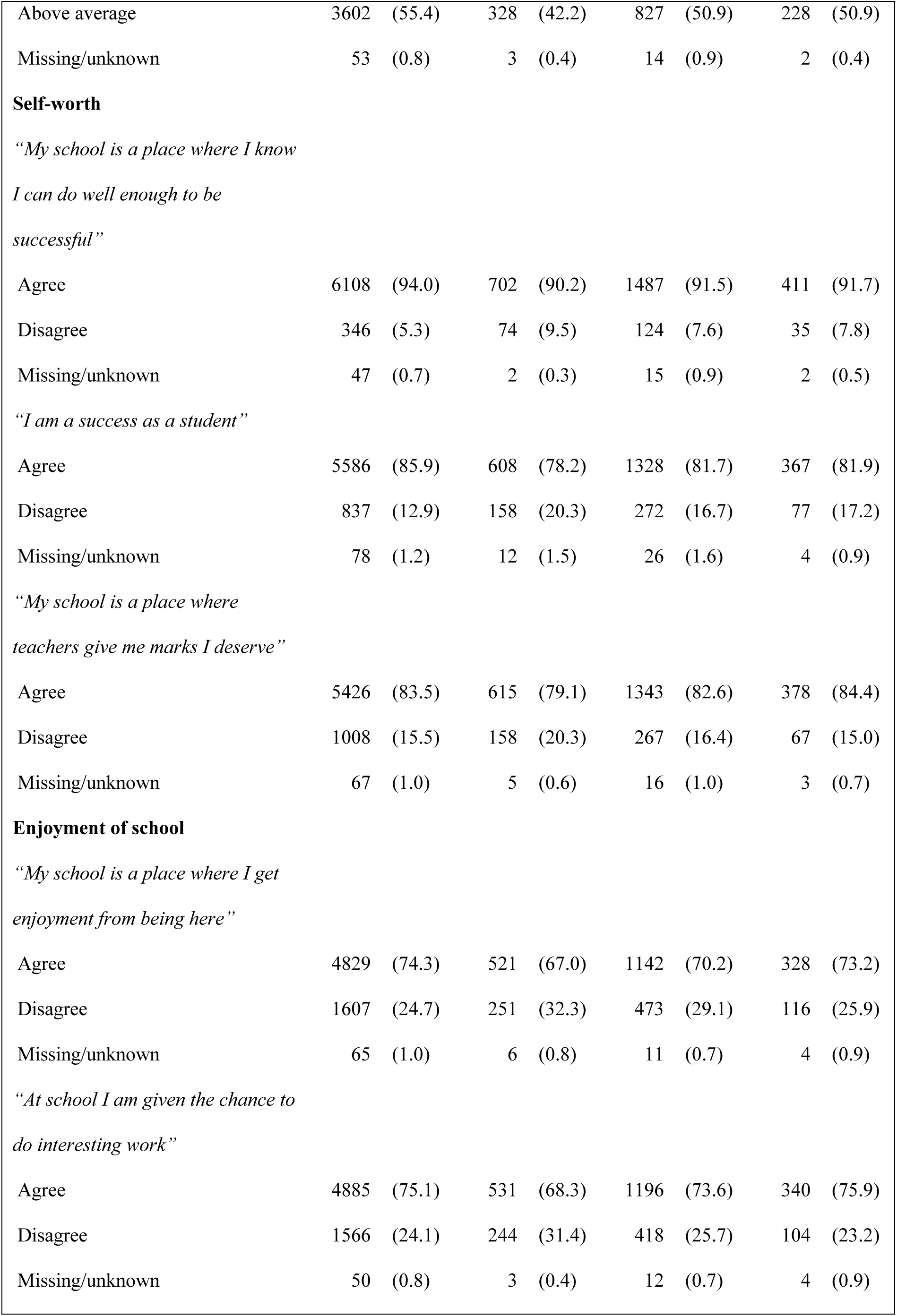
Summary statistics for binary and categorical predictors for model 2 trajectory groups. Percentages may not equal 100 due to rounding.

[Insert Table 6 about here]
| Characteristic | Assigned Trajectory Group<sup>^</sup> |  |  |  |  |  |  |  |
| --- | --- | --- | --- | --- | --- | --- | --- | --- |
|  | **Regular**<br>**weekly**<br>**exerciser**<br>**(n=6501)** |  | **Infrequent**<br>**exerciser**<br>**(n=778)** |  | **Decreasing**<br>**exerciser**<br>**(n=1626)** |  | **Increasing**<br>**exerciser**<br>**(n=448)** |  |
|  | Freq | (%) | Freq | (%) | Freq | (%) | Freq | (%) |
| **Gender** |  |  |  |  |  |  |  |  |
| Male | 3372 | (51.9) | 244 | (31.4) | 686 | (42.2) | 159 | (35.5) |
| Female | 3129 | (48.1) | 534 | (68.6) | 940 | (57.8) | 289 | (64.5) |
| **Indigenous Status** |  |  |  |  |  |  |  |  |
| Non-indigenous | 6141 | (94.5) | 723 | (92.9) | 1540 | (94.7) | 431 | (96.2) |
| Indigenous | 360 | (5.5) | 55 | (7.1) | 86 | (5.3) | 17 | (3.8) |
| **Self-efficacy** |  |  |  |  |  |  |  |  |
| *“Compared with most students in your year level, how well are you doing in your subjects overall?”* |  |  |  |  |  |  |  |  |
| Below average | 241 | (3.7) | 57 | (7.3) | 79 | (4.9) | 24 | (5.4) |
| About average | 2605 | (40.1) | 390 | (50.1) | 706 | (43.4) | 194 | (43.3) |

|  |  |  |  |  |  |  |  |  |
| --- | --- | --- | --- | --- | --- | --- | --- | --- |
| Above average | 3602 | (55.4) | 328 | (42.2) | 827 | (50.9) | 228 | (50.9) |
| Missing/unknown | 53 | (0.8) | 3 | (0.4) | 14 | (0.9) | 2 | (0.4) |
| **Self-worth** |  |  |  |  |  |  |  |  |
| *“My school is a place where I know* |  |  |  |  |  |  |  |  |
| *I can do well enough to be* |  |  |  |  |  |  |  |  |
| *successful”* |  |  |  |  |  |  |  |  |
| Agree | 6108 | (94.0) | 702 | (90.2) | 1487 | (91.5) | 411 | (91.7) |
| Disagree | 346 | (5.3) | 74 | (9.5) | 124 | (7.6) | 35 | (7.8) |
| Missing/unknown | 47 | (0.7) | 2 | (0.3) | 15 | (0.9) | 2 | (0.5) |
| *“I am a success as a student”* |  |  |  |  |  |  |  |  |
| Agree | 5586 | (85.9) | 608 | (78.2) | 1328 | (81.7) | 367 | (81.9) |
| Disagree | 837 | (12.9) | 158 | (20.3) | 272 | (16.7) | 77 | (17.2) |
| Missing/unknown | 78 | (1.2) | 12 | (1.5) | 26 | (1.6) | 4 | (0.9) |
| *“My school is a place where* |  |  |  |  |  |  |  |  |
| *teachers give me marks I deserve”* |  |  |  |  |  |  |  |  |
| Agree | 5426 | (83.5) | 615 | (79.1) | 1343 | (82.6) | 378 | (84.4) |
| Disagree | 1008 | (15.5) | 158 | (20.3) | 267 | (16.4) | 67 | (15.0) |
| Missing/unknown | 67 | (1.0) | 5 | (0.6) | 16 | (1.0) | 3 | (0.7) |
| **Enjoyment of school** |  |  |  |  |  |  |  |  |
| *“My school is a place where I get* |  |  |  |  |  |  |  |  |
| *enjoyment from being here”* |  |  |  |  |  |  |  |  |
| Agree | 4829 | (74.3) | 521 | (67.0) | 1142 | (70.2) | 328 | (73.2) |
| Disagree | 1607 | (24.7) | 251 | (32.3) | 473 | (29.1) | 116 | (25.9) |
| Missing/unknown | 65 | (1.0) | 6 | (0.8) | 11 | (0.7) | 4 | (0.9) |
| *“At school I am given the chance to* |  |  |  |  |  |  |  |  |
| *do interesting work”* |  |  |  |  |  |  |  |  |
| Agree | 4885 | (75.1) | 531 | (68.3) | 1196 | (73.6) | 340 | (75.9) |
| Disagree | 1566 | (24.1) | 244 | (31.4) | 418 | (25.7) | 104 | (23.2) |
| Missing/unknown | 50 | (0.8) | 3 | (0.4) | 12 | (0.7) | 4 | (0.9) |

Relative to the *regular weekly* exercise group, below average self-efficacy (versus above-average self-efficacy) was a risk factor for membership in *infrequent exercisers* and *declining-exerciser* groups (aRRR for *infrequent exercisers* =1.85, 95% CI 1.26, 2.72; p=0.002; aRRR for *declining exercisers* =1.45, 95% CI 1.07, 1.97; p=0.017) (Fig 3, A and B). Higher paternal ISEI was protective against membership in the *infrequent exercisers* and *declining-exerciser* groups (for every 10 units in ISEI score, aRRR for *infrequent exercisers*=0.92; 95% CI 0.88, 0.97; p=0.001; aRRR for *declining exercisers* =0.95, 95% CI 0.92, 0.98; p=0.002) (Figs 6 - 8, Table 6).

Young people with higher academic literacy scores were more likely to fall into either the *decreasing exerciser* or *increasing exerciser* groups relative to the *regular weekly* exercise group (for every 1000 units, aRRR for *decreasing exerciser* =1.11; 95% CI 1.05, 1.18; p=0.001; aRRR for *increasing exerciser* =1.33, 95% CI 1.20, 1.47; p<.0001) (Fig 2, b and c). Indigenous status, measures of self-worth, enjoyment of school, or interest in schoolwork were not found to be significantly associated with trajectory group (Figs 6 - 8, Tables 5 and 6).

**Legend Figs 6 - 8.**
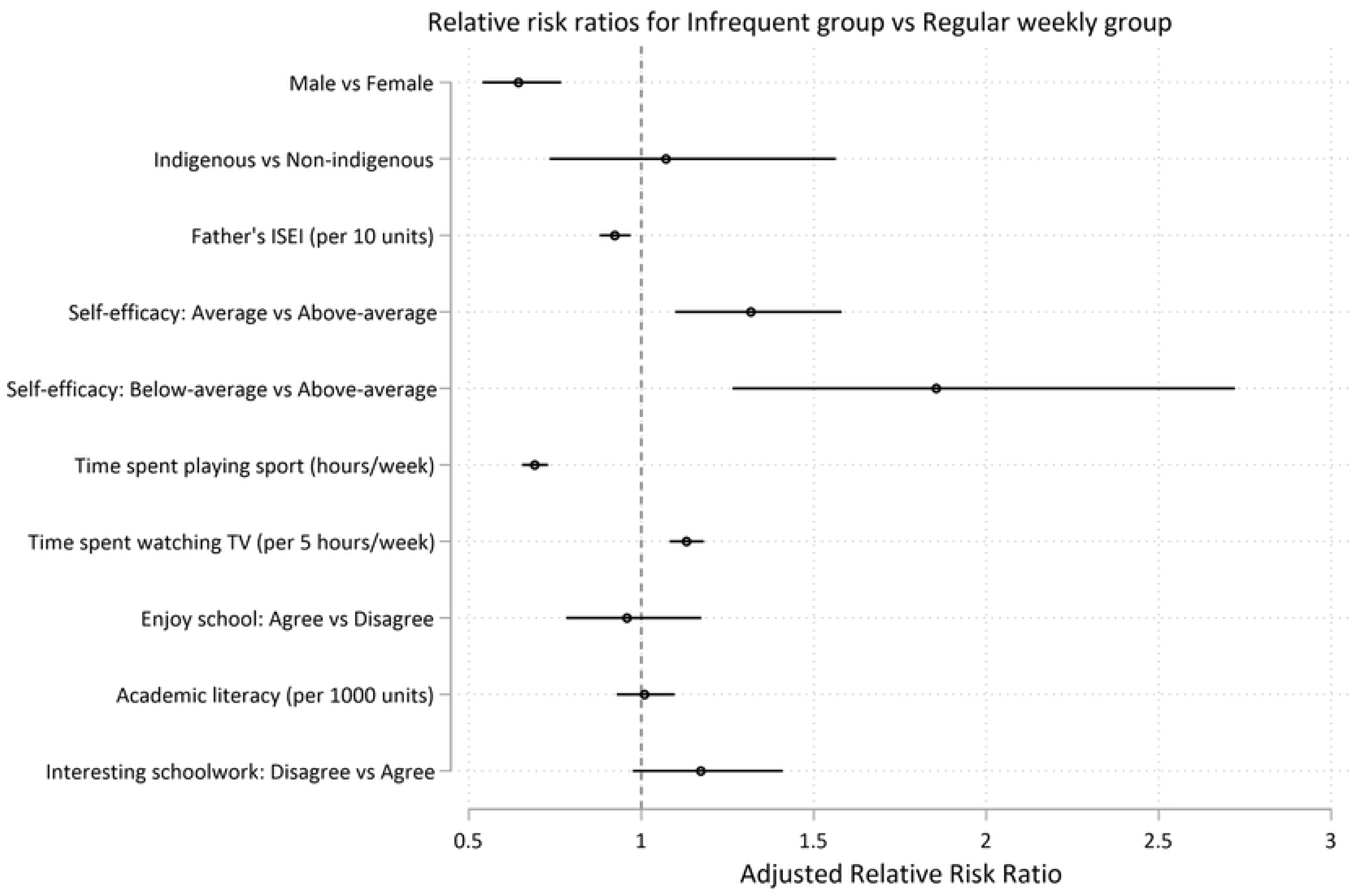
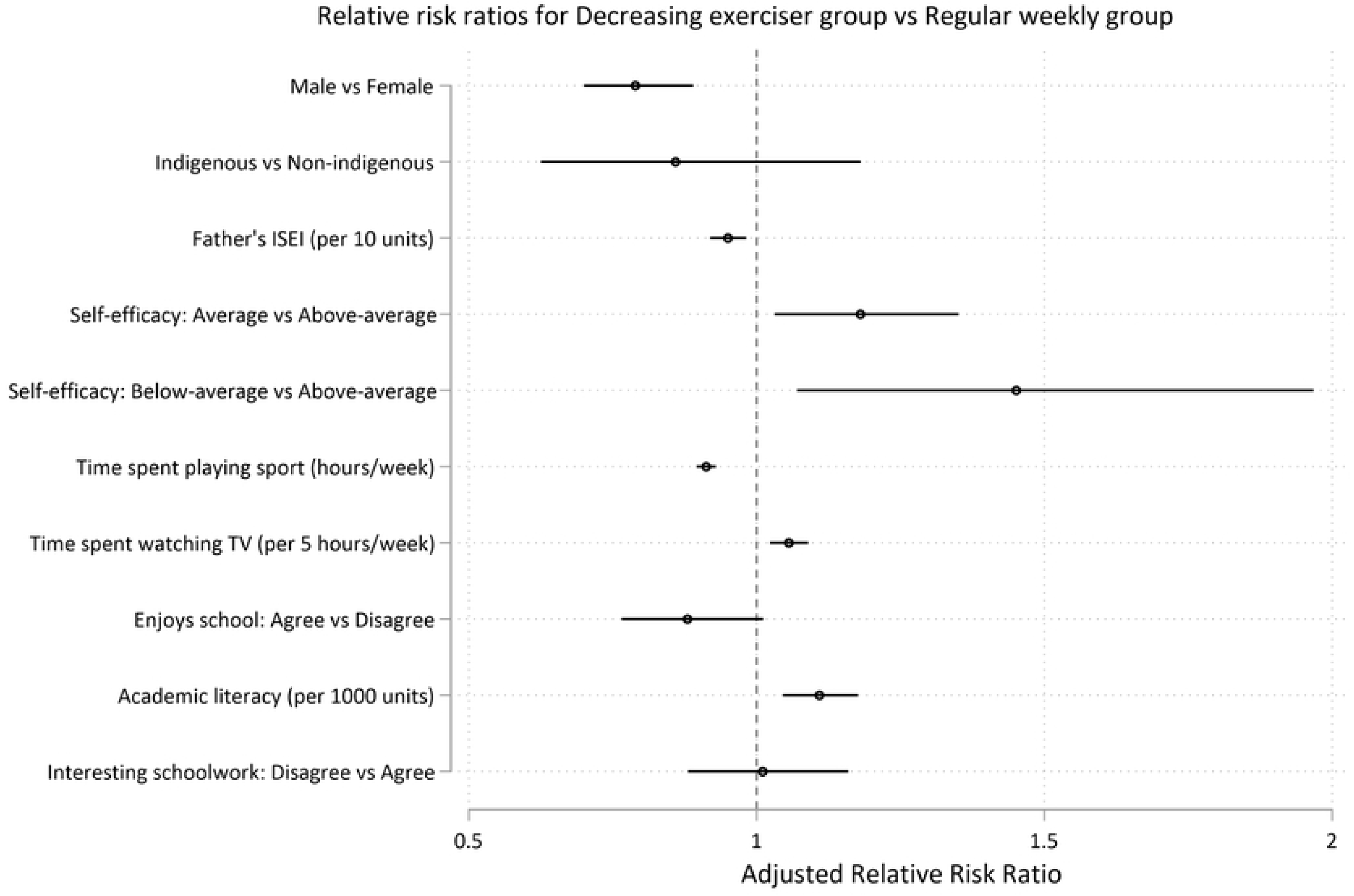
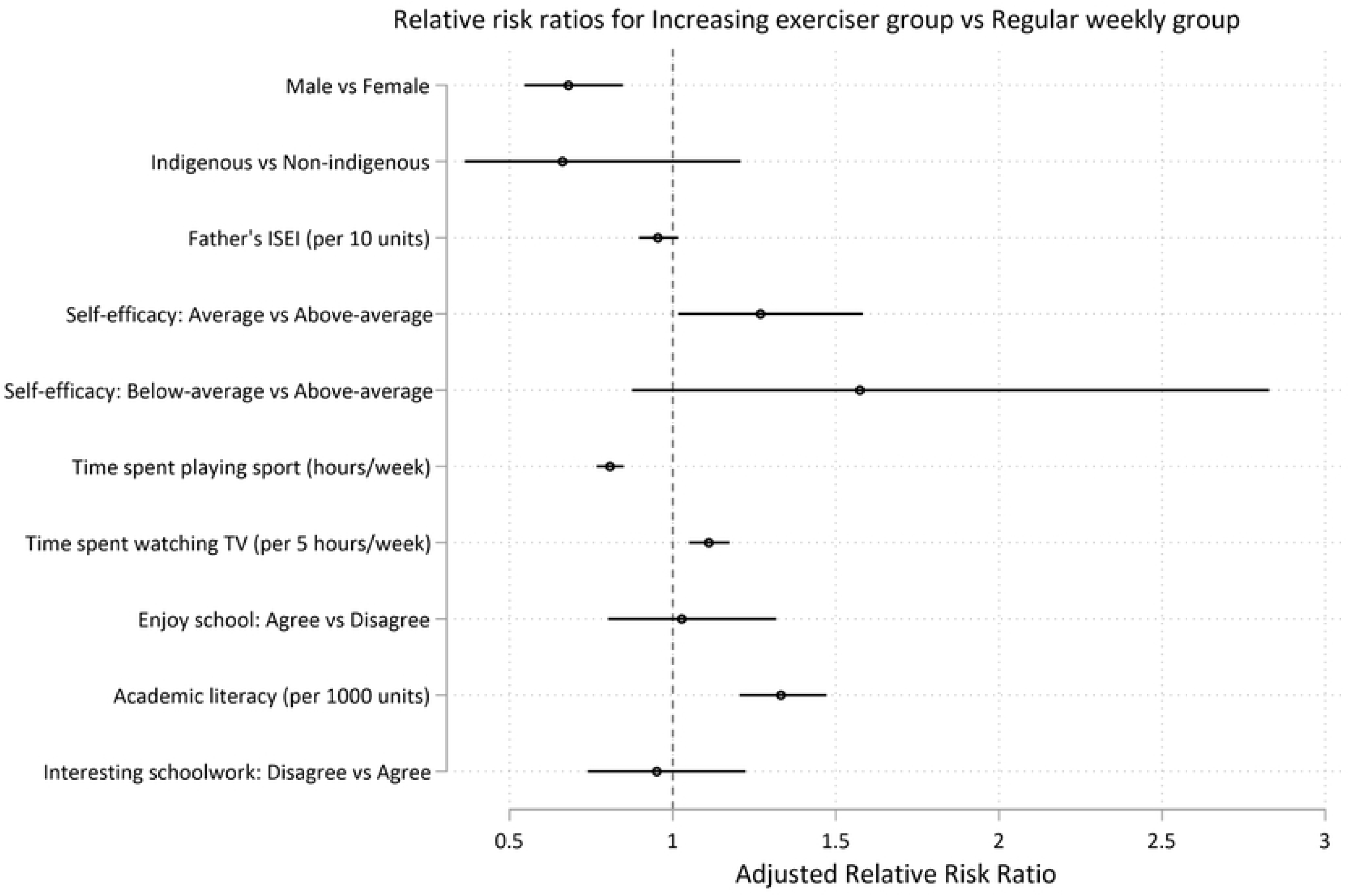
Predictors at age 15 for long-term recreational physical exercise participation between the ages of 16 and 24. Risks for insufficient (less than once weekly) recreational exercise (Model 2). Risks are shown as adjusted Relative Risk Ratios (aRRRs) for falling into the infrequent exerciser trajectory (Fig 6), the decreasing exerciser trajectory (Fig 7) and the increasing exerciser trajectory (Fig 8), compared to the risk of following the regular weekly exercise trajectory. ISEI: International Socio-Economic Index of Occupational Status.

### 3.3 Young people who maintain daily recreational exercise between age 16 and 24 report better general health at age 25 than those who don’t exercise daily; no additional advantages on measured indicators of mental health and vocational attainment were identified

Guideline-adherent exerciser group membership was positively associated with self-reported general health at age 25, and this association remained after the adjustment for possible confounders of gender, ethnicity, participants’ ISEI score, and the predictors of trajectory group membership.

Compared to the *guideline* exerciser group, the *never guideline*, *guideline dropouts*, and *towards guideline* groups had significantly higher odds of poorer self-reported general health (results reported as adjusted Odds Ratio (aOR) of lower vs higher self-reported general health, 95% CI; p); *guideline* vs *never guideline* =2.24, 95% CI 1.85, 2.73; p<.0001; *guideline* vs *guideline dropouts* =1.91, 95% CI 1.45, 2.51; p<.0001; *guideline* vs *towards guideline* =1.56, 95% CI 1.29, 1.90; p<.0001)(Fig 9). There were no associations between trajectory groups (according to probability of daily exercise over time) and mental illness, life satisfaction at age 25 (happiness with the future or happiness with life overall), year 12 completion, the attainment of a post school qualification, or participation in the labour force (Figs 10 - 15).

**Legend Figs 9 - 15.**
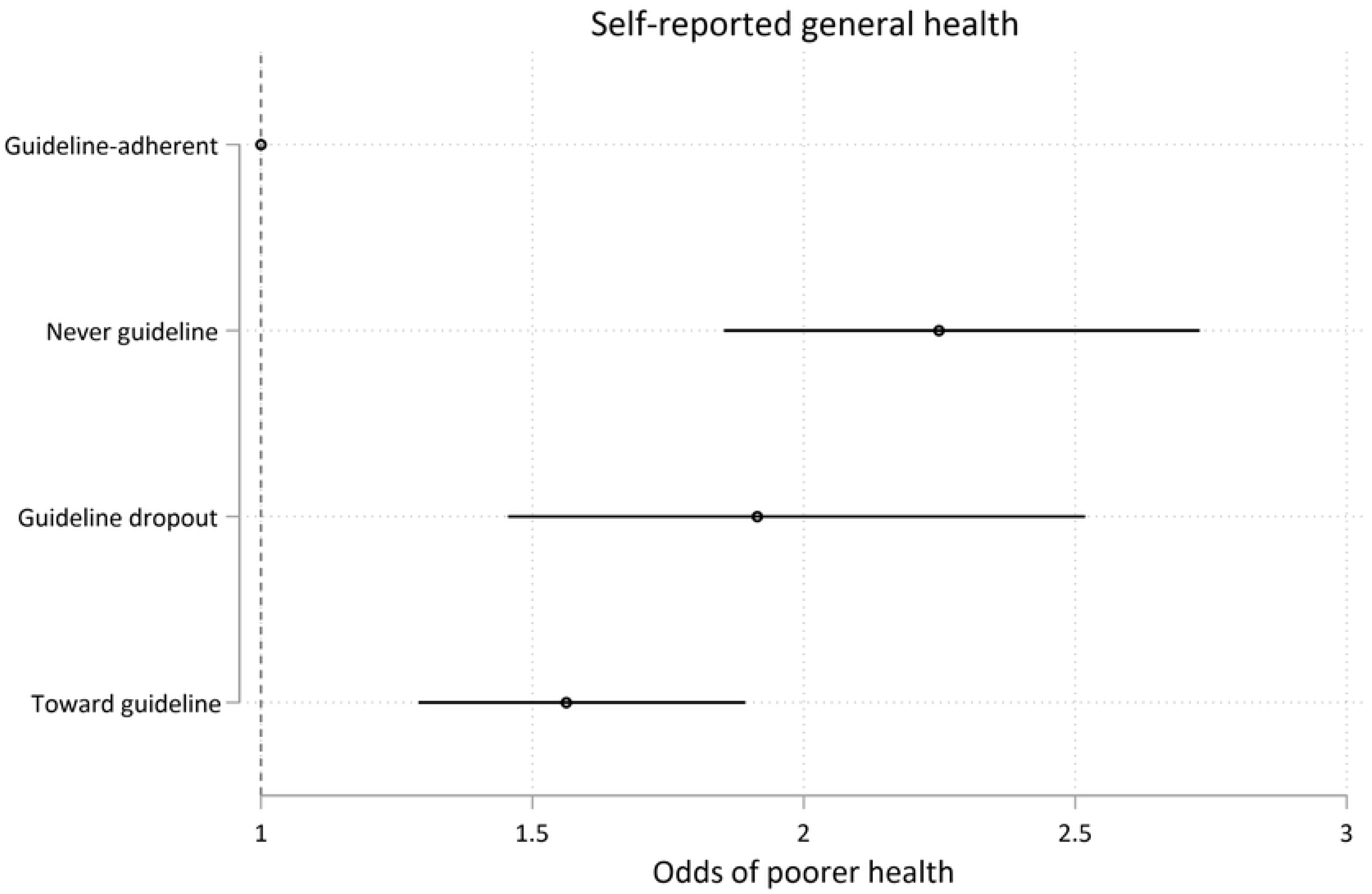
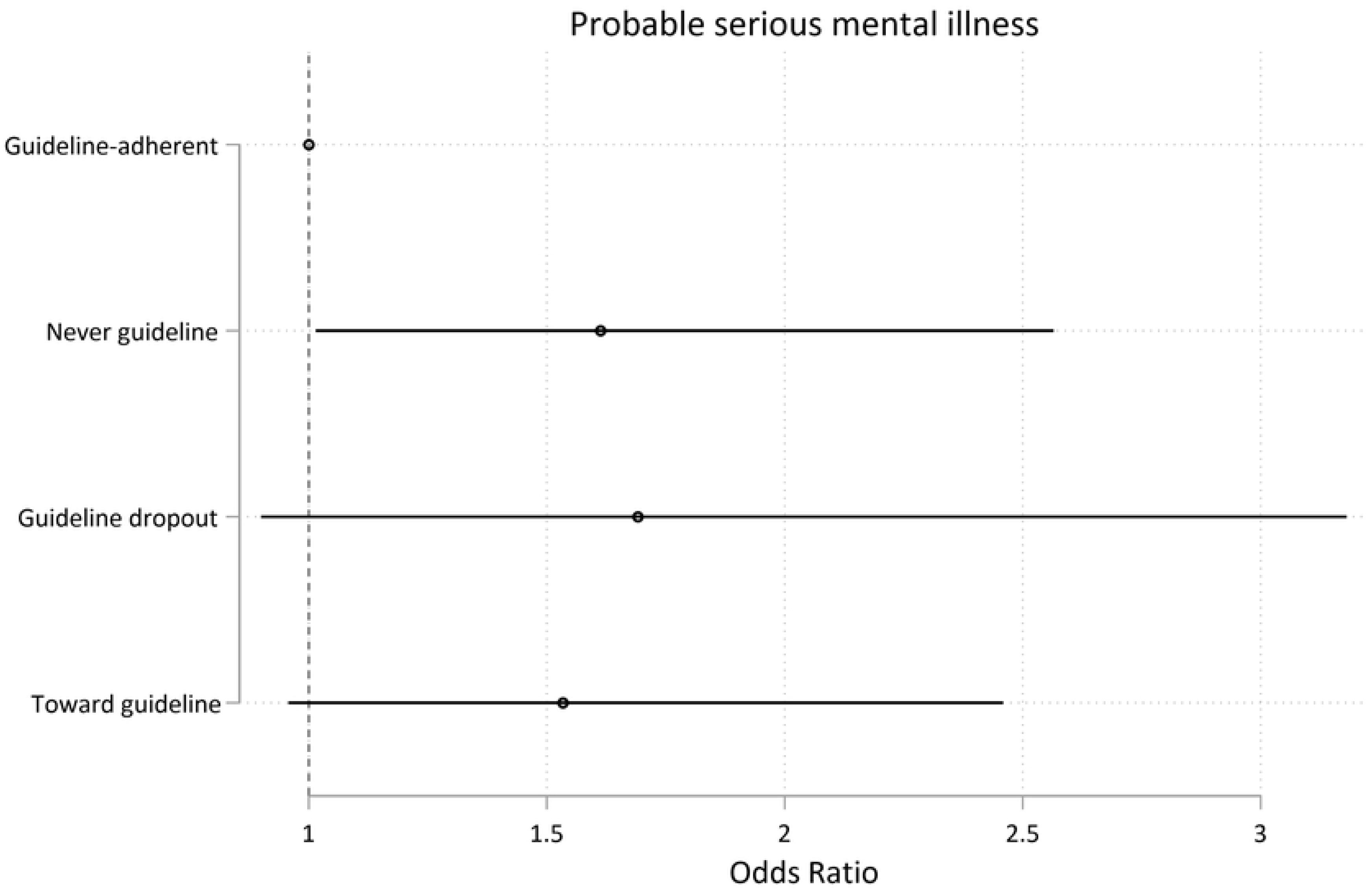
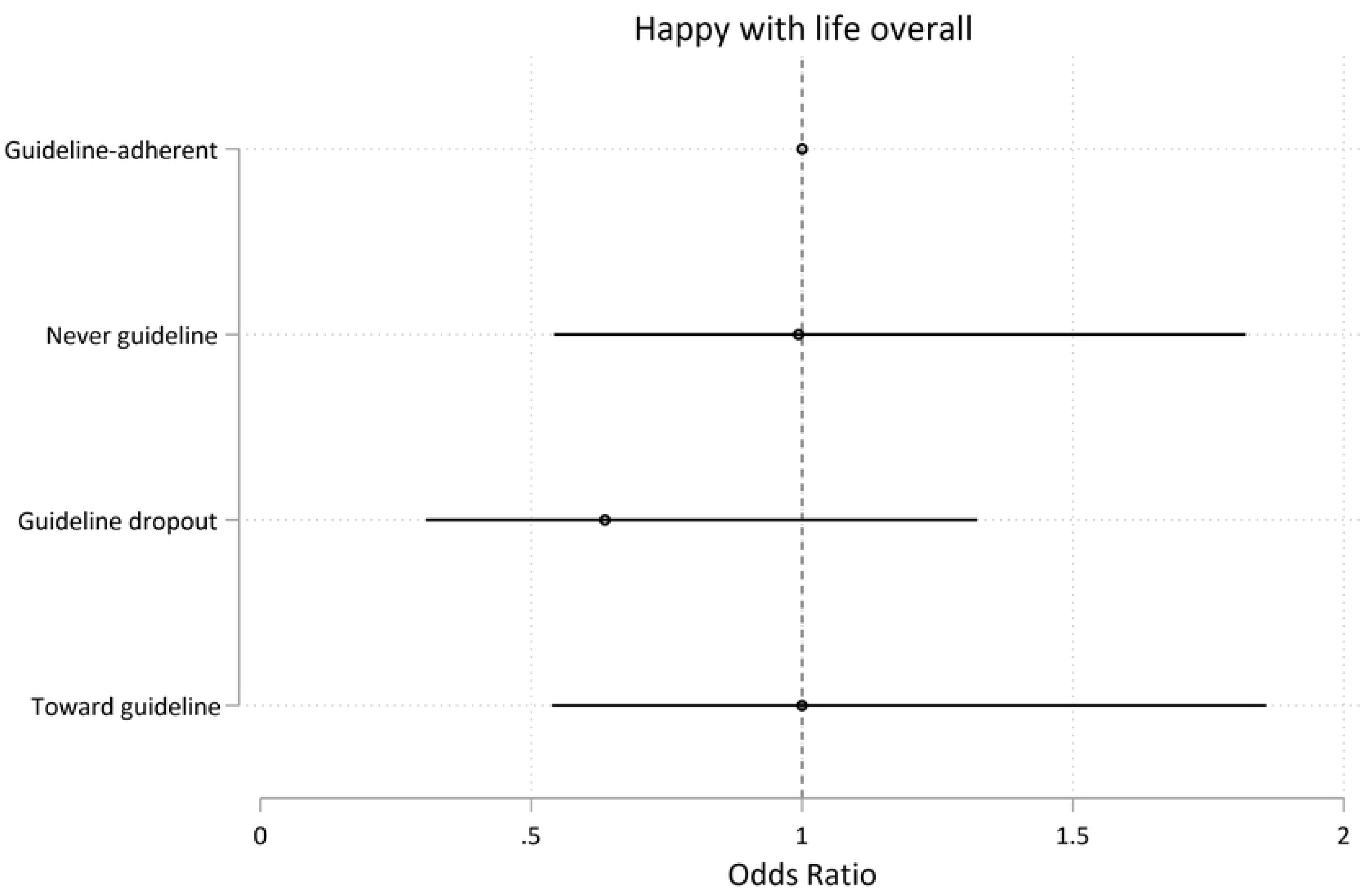
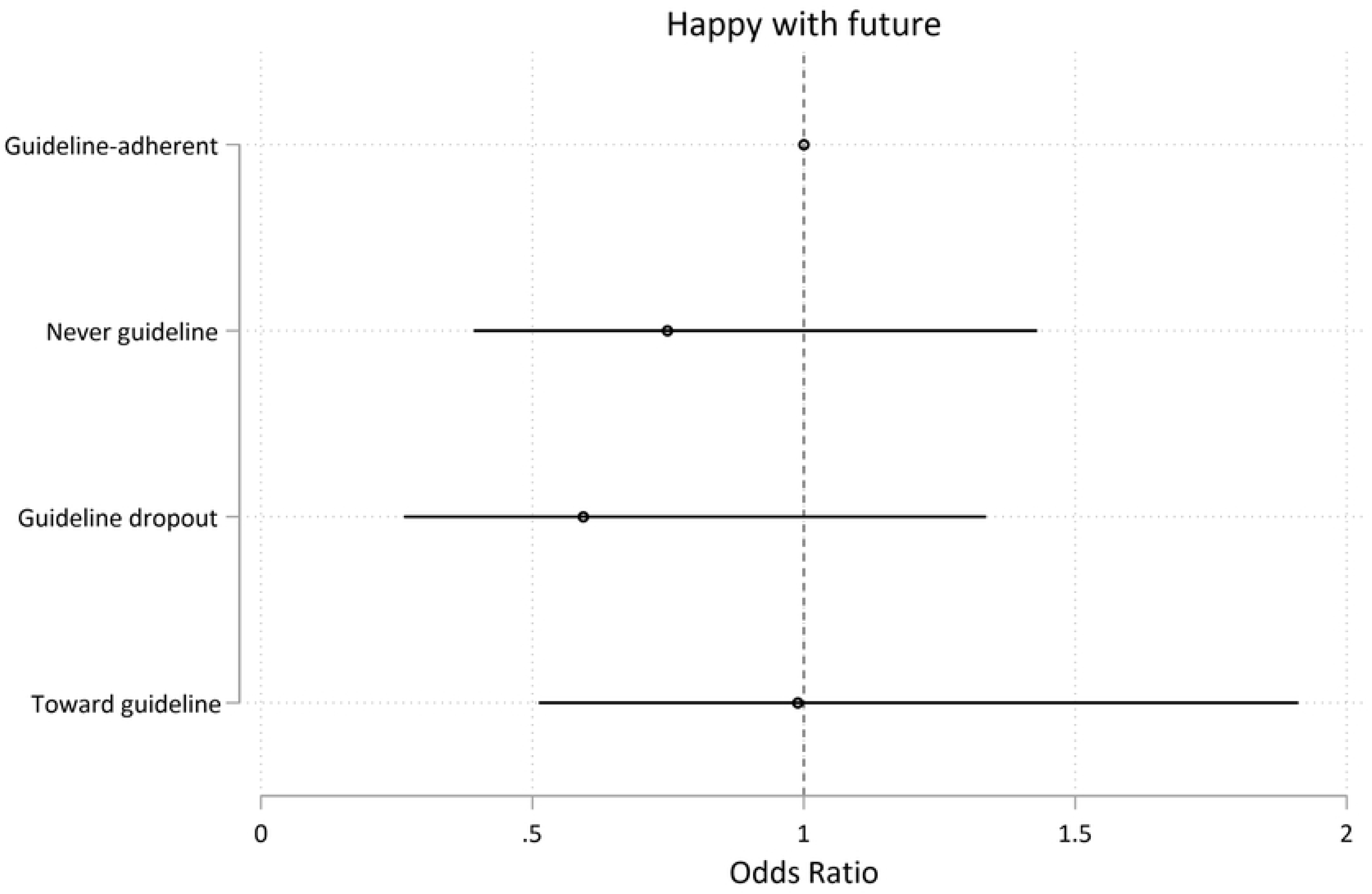
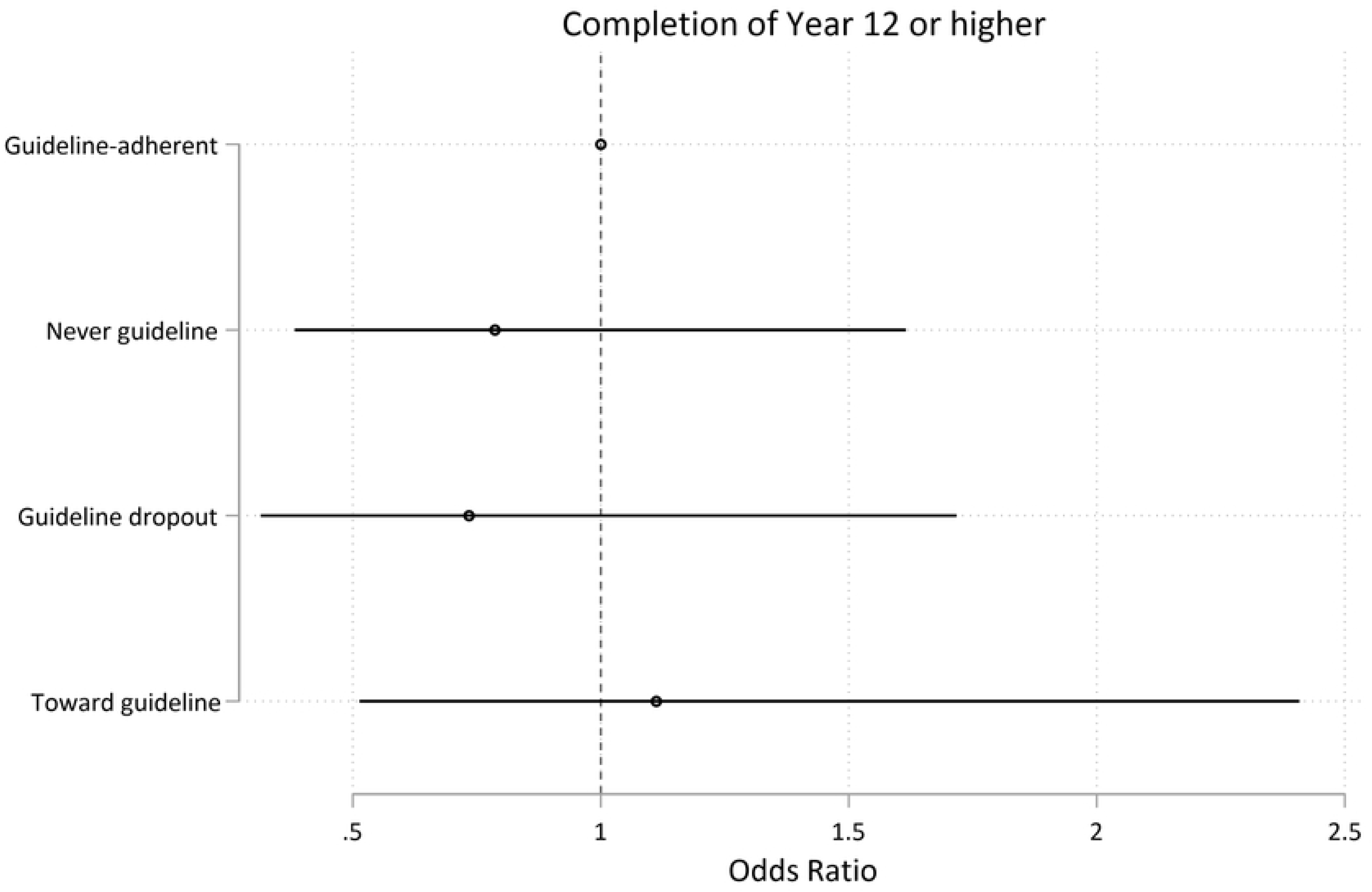
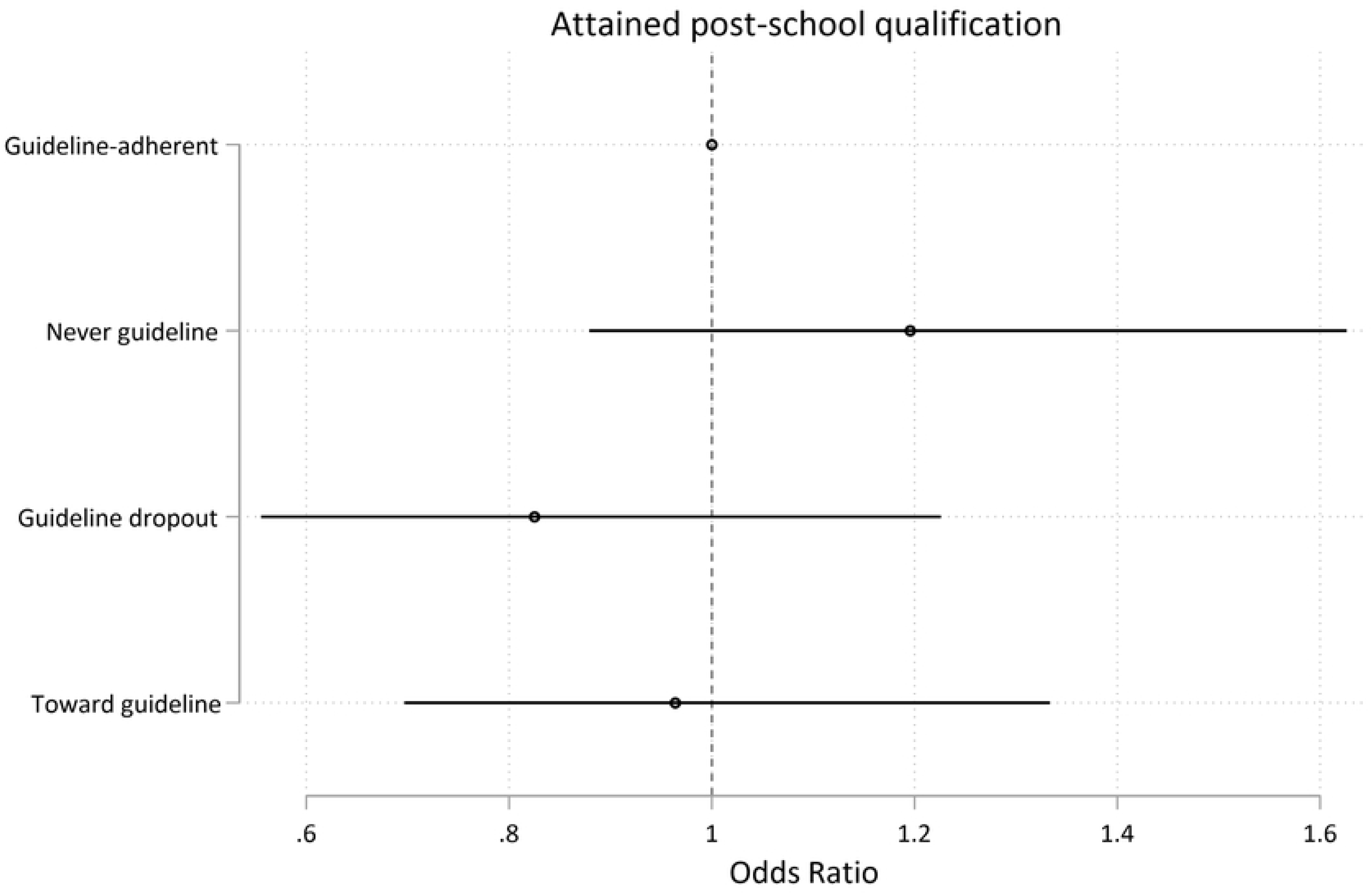
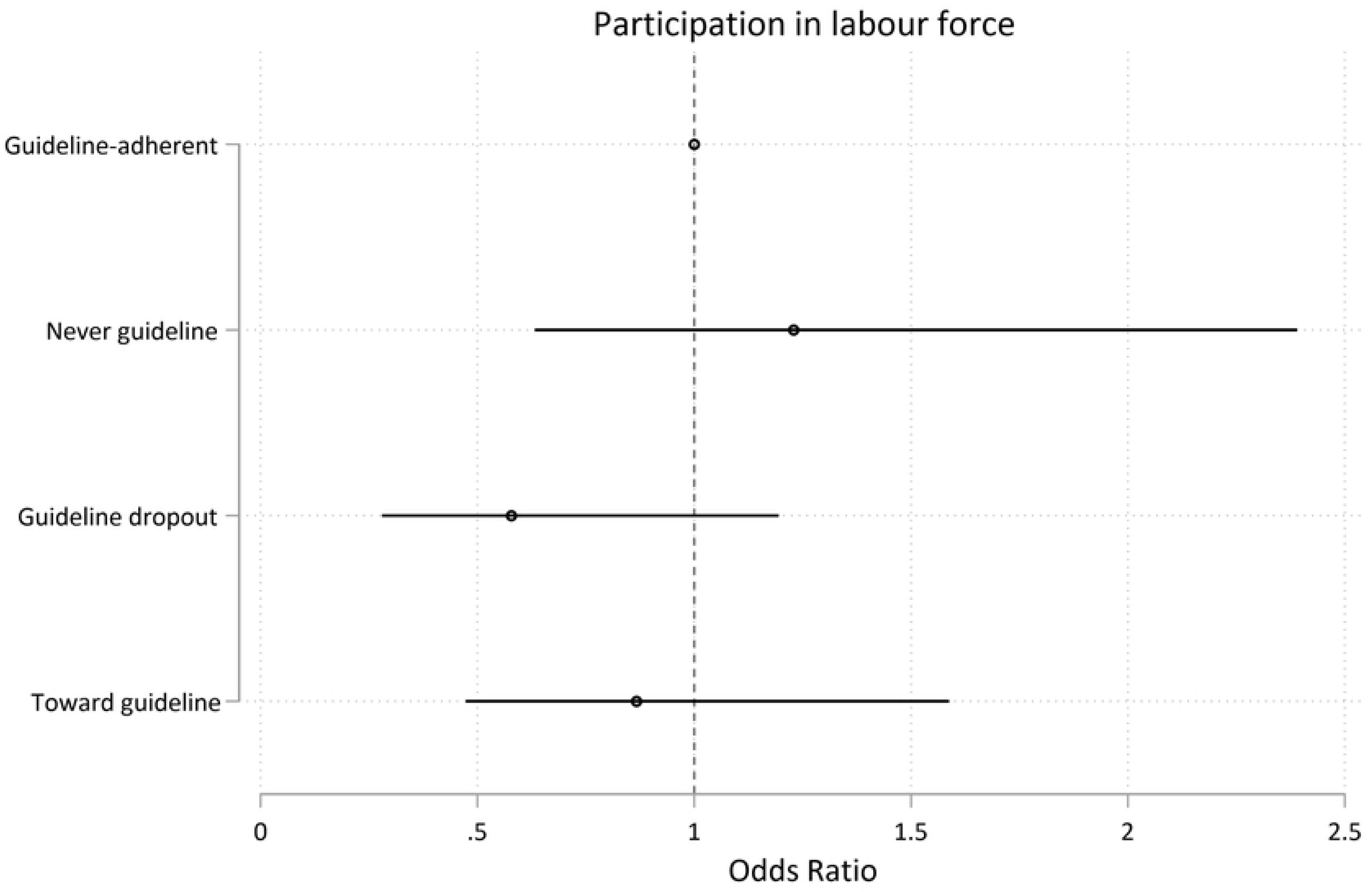
Model 1 trajectory group associations with health-, mental health-, and educational outcomes at age 25. Associations expressed as adjusted odds ratios (ORs) describing the odds of experiencing each outcome of interest for participants who undertake less than daily recreational physical exercise (the *never guideline, guideline dropouts,* and *towards guideline* exercise trajectories) vs participants undertaking the recommended level of recreational physical exercise (*guideline* exercisers). Less than *guideline* levels of recreational exercise had a) poorer self-reported general health (Fig 9). There were no associations between less than *guideline* levels of recreational exercise and Fig 10) mental illness Fig 11) overall satisfaction with life; Fig 12) satisfaction with the future; Fig 13) completing high school year 12, or higher; Fig 14) attaining any post-school qualification; Fig 15) participation in the labour force.

**Table 7.** Summary statistics of Model 1 trajectory group associations with outcomes at age 25.

| Outcome | Guideline-adherent |  | Never guideline |  | Guideline drop-out |  | Towards guideline |  |
| --- | --- | --- | --- | --- | --- | --- | --- | --- |
|  | Freq | (%) | Freq | (%) | Freq | (%) | Freq | (%) |
| **Psychological distress** |  |  |  |  |  |  |  |  |
| **(Kessler-6)** |  |  |  |  |  |  |  |  |
| Lower risk of mental illness | 543/567 | (95.8) | 1307/1420 | (92.0) | 267/288 | (92.7) | 939/1020 | (92.1) |
| Greater risk of mental illness | 24/567 | (4.2) | 113/1420 | (8.0) | 21/288 | (7.3) | 81/1020 | (7.9) |
| **Self-reported general health** |  |  |  |  |  |  |  |  |
| Excellent | 169/574 | (29.4) | 184/1432 | (12.9) | 54/291 | (18.6) | 217/1030 | (21.1) |
| Very good | 247/574 | (43.0) | 535/1432 | (37.4) | 110/291 | (37.8) | 367/1030 | (35.6) |
| Good | 115/574 | (20.0) | 503/1432 | (35.5) | 91/291 | (31.3) | 329/1030 | (31.9) |
| Fair | 30/574 | (5.2) | 170/1432 | (11.9) | 31/291 | (10.7) | 96/1030 | (9.3) |
| Poor | 13/574 | (2.3) | 35/1432 | (2.4) | 5/291 | (1.7) | 21/1030 | (2.0) |
| **Life satisfaction: happy with life as a whole** |  |  |  |  |  |  |  |  |
| Happy | 553/573 | (96.5) | 1350/1416 | (95.3) | 270/287 | (94.1) | 969/1014 | (95.6) |
| Unhappy | 20/573 | (3.5) | 66/1416 | (4.7) | 17/287 | (5.9) | 45/1014 | (4.4) |
| **Life satisfaction: happy with the future** |  |  |  |  |  |  |  |  |
| Happy | 550/565 | (97.4) | 1315/1381 | (95.2) | 266/280 | (95.0) | 961/995 | (96.6) |
| Unhappy | 15/565 | (2.7) | 66/1381 | (4.8) | 14/280 | (5.0) | 34/995 | (3.4) |
| **Completion of high school (Year 12 or Cert II)** |  |  |  |  |  |  |  |  |
| Completed Year 12/Cert II | 558/575 | (97.0) | 1391/1438 | (96.7) | 281/292 | (96.2) | 1004/1038 | (96.7) |
| Did not complete | 17/575 | (3.0) | 47/1438 | (3.3) | 11/292 | (3.8) | 34/1038 | (3.3) |
|  | Freq | (%) | Freq | (%) | Freq | (%) | Freq | (%) |
| **Completion of any post-school qualification** |  |  |  |  |  |  |  |  |
| Yes | 491/575 | (85.4) | 1247/1438 | (86.7) | 240/292 | (82.2) | 871/1038 | (83.9) |
| No | 84/575 | (14.6) | 191/1438 | (13.3) | 52/292 | (17.8) | 167/1038 | (16.1) |
| **Participation in the labour force** |  |  |  |  |  |  |  |  |
| Employed | 535/552 | (96.9) | 1302/1340 | (97.2) | 256/270 | (94.8) | 945/983 | (96.1) |
| Unemployed | 17/552 | (3.1) | 38/1340 | (2.8) | 14/270 | (5.2) | 38/983 | (3.9) |

### 3.4 Young people with insufficient levels of recreational exercise between age 16 and 24 are at higher risk of adverse health-and mental health-outcomes at age 25

Membership in less-than weekly exercise trajectory groups were negatively associated with measures of general health, psychological distress, and life satisfaction at age 25, and associations persisted after adjusting for the potentially confounding effects of gender, ethnicity, participants’ ISEI score, and other identified predictor variables (Figs 16 - 19, Table 5). The associations between trajectory-group membership and the attainment of school-and post-school qualifications, and with participation in the labour force did not persist following adjustment. (Figs 20 - 22, Table 5).

Compared to the *regular weekly* exercise group, *infrequent exercisers, declining exercisers, and increasing-exercisers* had significantly higher odds of poorer self-reported general health (results reported as adjusted Odds Ratio (aOR), 95% CI; p); for *infrequent exercisers* =2.57, 95% CI 2.02, 3.28; p<.0001; for *declining exercisers* =1.97, 95% CI 1.66, 2.34; p<.0001; aOR for *increasing exercisers* =1.81, 95% CI 1.46, 2.24; p<.0001)(Fig 16), and had higher odds of elevated K6 scores suggesting probable mental illness (aOR for *infrequent exercisers*=2.42, 95% CI 1.55, 3.78; p<.0001; aOR for *declining exercisers* =2.19, 95% CI 1.64, 2.93; p<.0001; aOR for *increasing exercisers* =1.59, 95% CI 1.03, 2.47; p=0.036)(Fig 17). Young people in the *infrequent exercisers, declining exercisers*, and *increasing exercisers* groups were less likely to feel happy about the future; aOR for *infrequent exercisers* =0.25, 95% CI 0.15, 0.42; p<.0001; aOR for *declining exercisers* =0.51, 95% CI 0.33, 0.79; p=0.002; aOR for *increasing exercisers* =0.54, 95% CI 0.30, 0.97; p=0.041)(Fig 18), and the *infrequent exercisers* and *declining exercisers* groups were less likely to be satisfied with life overall (aOR for *infrequent exercisers* =0.33, 95% CI 0.20, 0.54; p<.0001; adjusted OR for *declining exercisers* =0.57, 95% CI 0.39, 0.84; p=0.004)(Fig 19).

For health-, mental health-, and life satisfaction measures we observed stepped decreases in the odds of adverse outcomes, with the *infrequent exercisers* group experiencing the highest disadvantage relative to *regular weekly* exercise, followed by *declining exercisers* and *increasing exercisers* (Figs 16 - 19). There is small variability in the total n available for each variable due to incomplete data (n range 3145-3343).

**Legend Figs 16 - 22.**
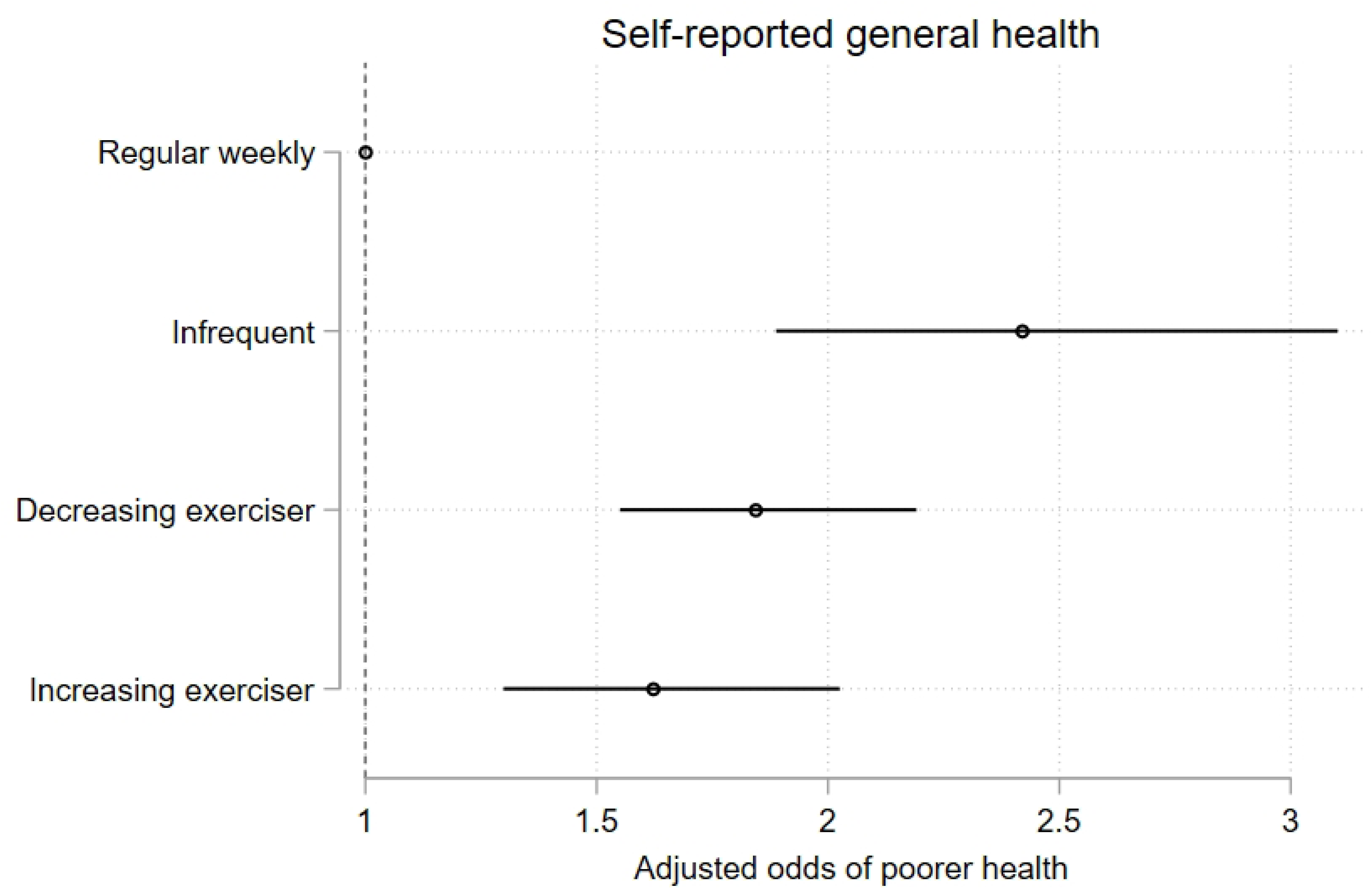
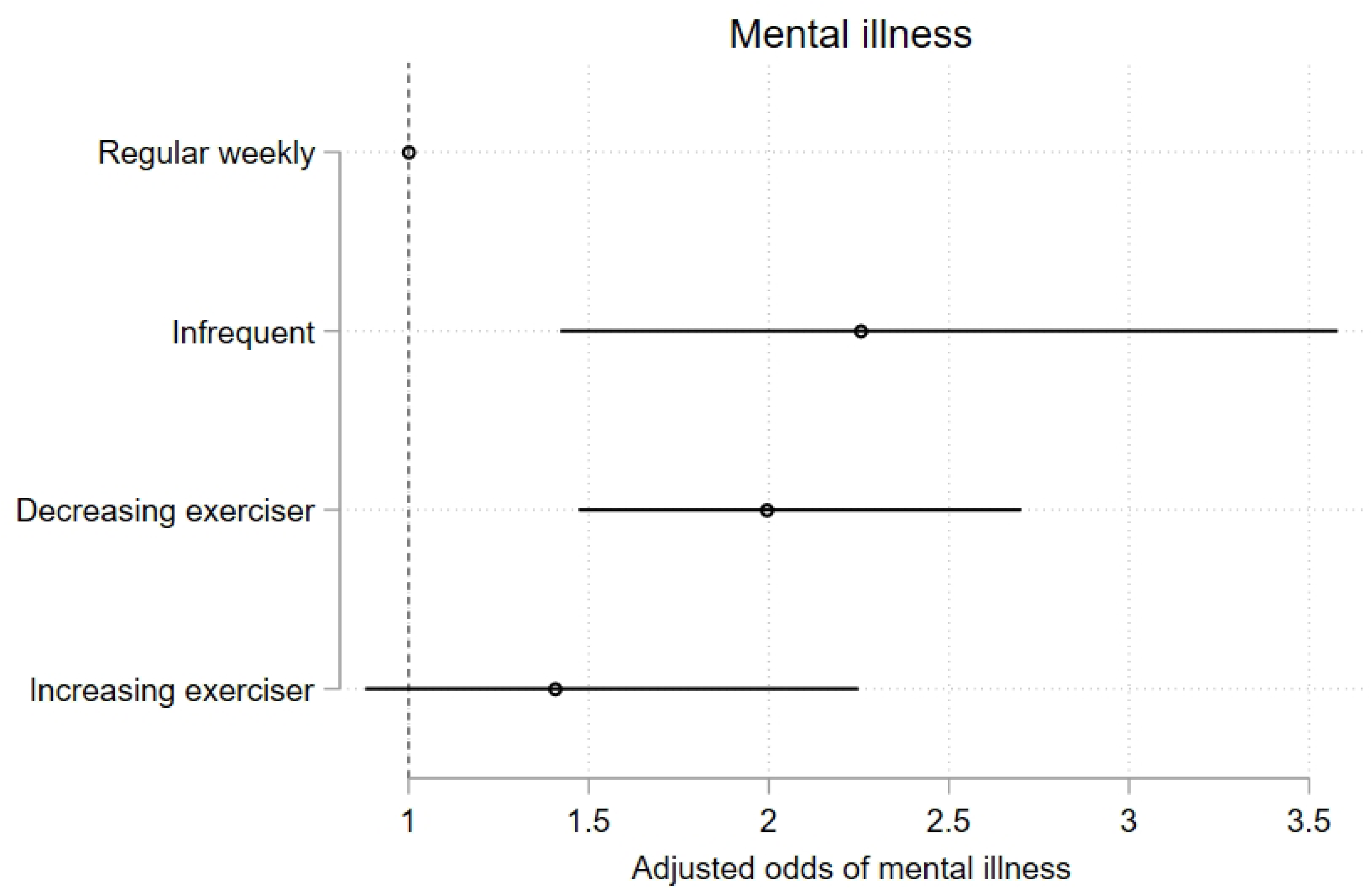
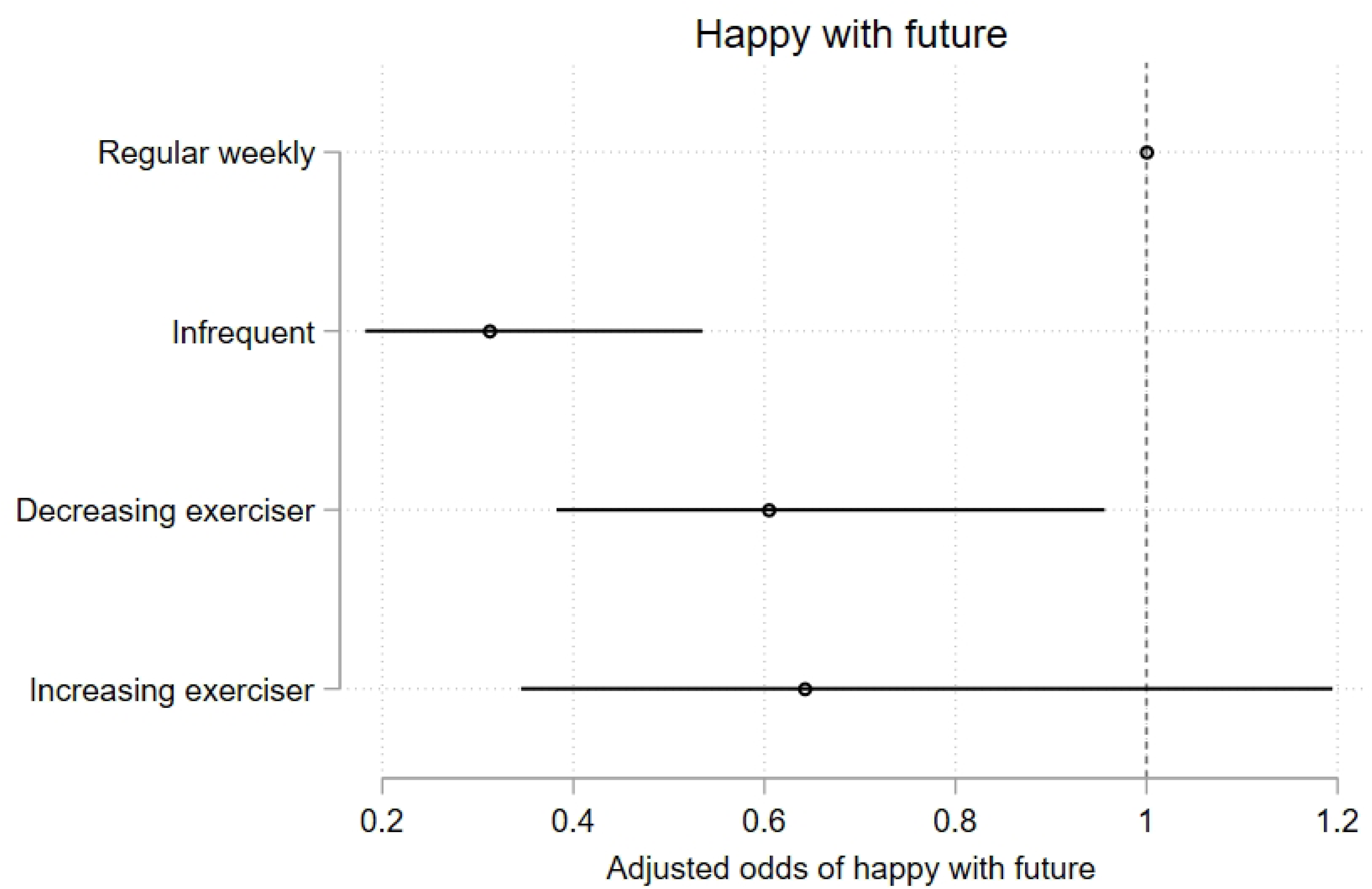
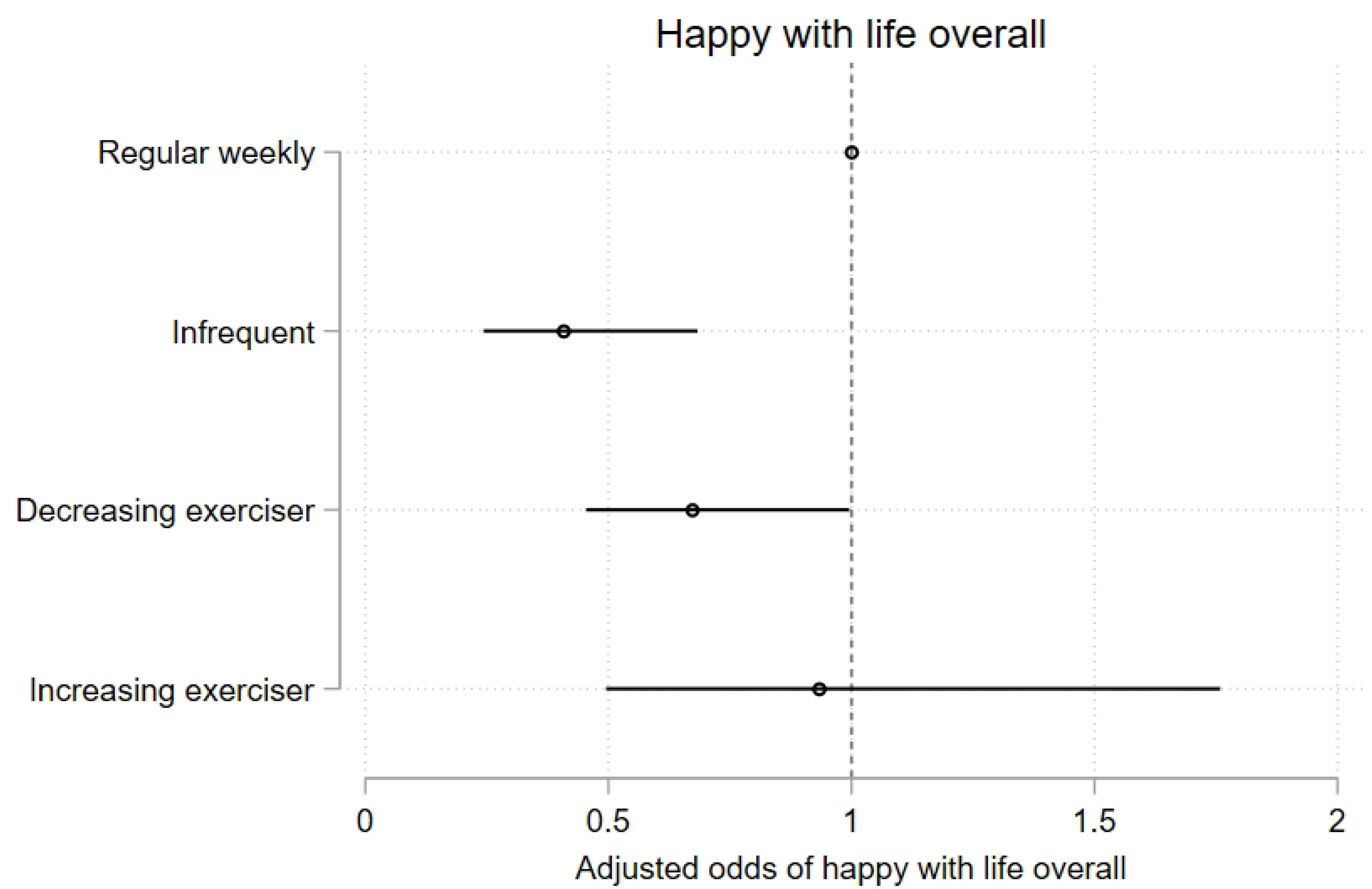
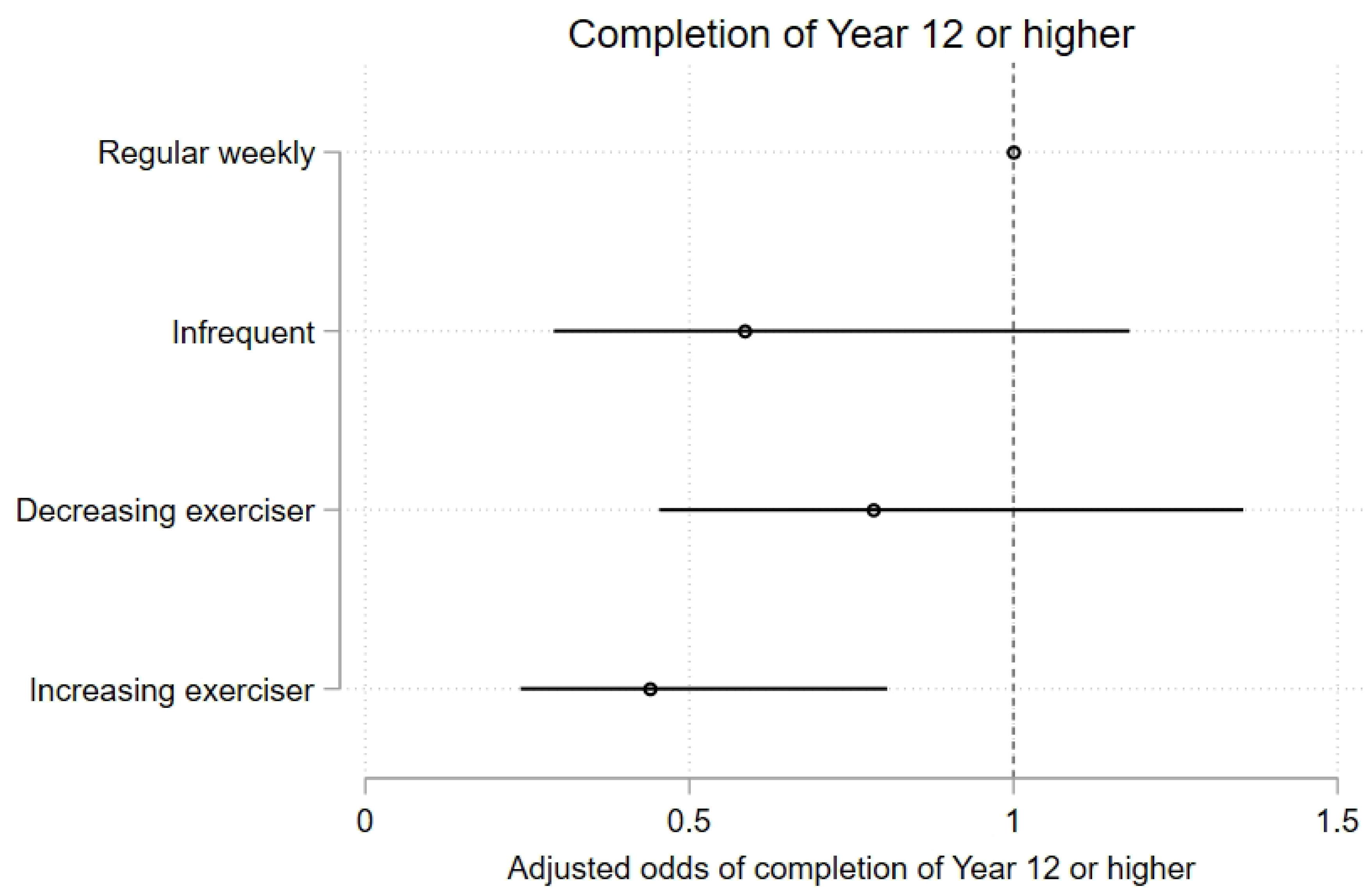
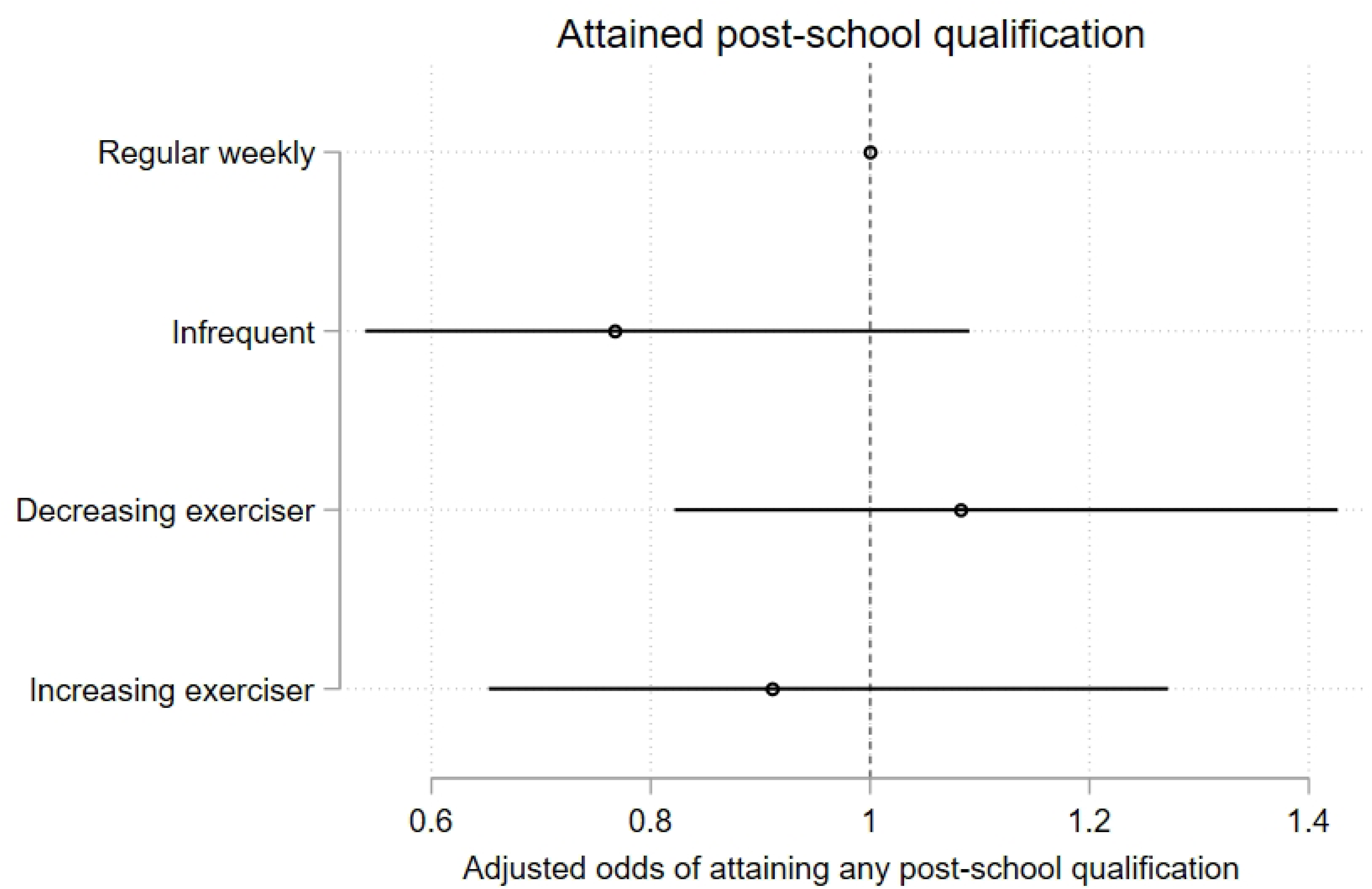
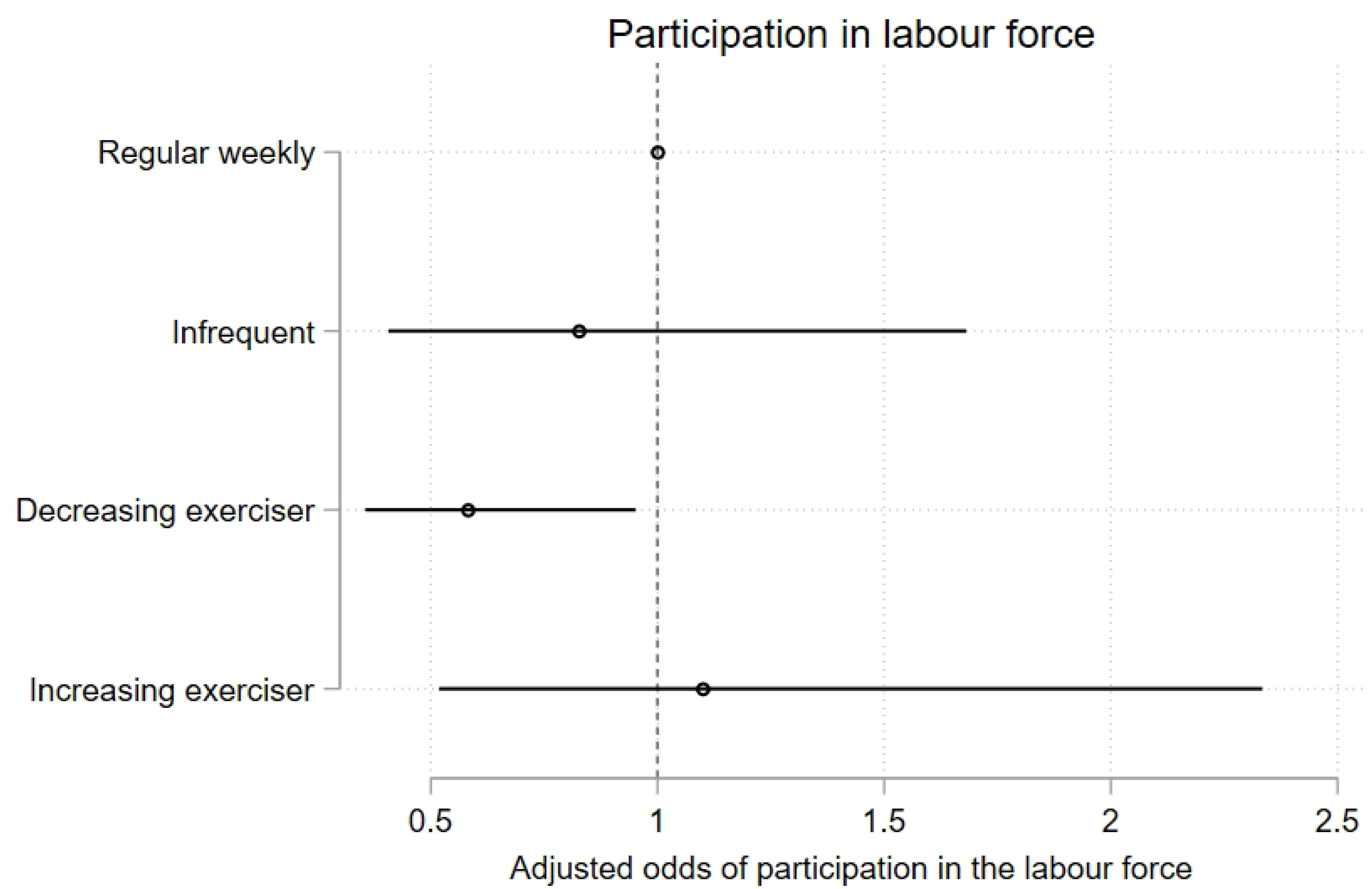
Model 2 trajectory group associations with health-, mental health-, and educational outcomes at age 25. Associations expressed as adjusted odds ratios (aORs) describing the odds of experiencing each outcome of interest for participants who undertake lower levels of recreational physical exercise (infrequent exercisers, decreasing exercisers, increasing exercisers) vs participants undertaking a higher level of recreational physical exercise (regular weekly exercisers). Outcomes are Fig 16) poorer general health; Fig 17) mental illness Fig 18) overall satisfaction with the future; Fig 19) overall satisfaction with life; Fig 20) completing high school year 12, or higher; Fig 21) attaining any post-school qualification; Fig 22) participation in the labour force.

**Table 8.** Summary statistics of Model 2 trajectory group associations with outcomes at age 25.

| Outcome | Regular weekly exerciser |  | Infrequent exerciser |  | Decreasing exerciser |  | Increasing exerciser |  |
| --- | --- | --- | --- | --- | --- | --- | --- | --- |
|  | Freq | (%) | Freq | (%) | Freq | (%) | Freq | (%) |
| **Psychological distress (Kessler-6)** |  |  |  |  |  |  |  |  |
| Lower risk of mental illness | 1900/2003 | (94.9) | 231/263 | (87.8) | 641/719 | (89.2) | 284/310 | (91.6) |
| Greater risk of mental illness | 103/2003 | (5.1) | 32/263 | (12.2) | 78/719 | (10.8) | 26/310 | (8.4) |
| **Self-reported general health** |  |  |  |  |  |  |  |  |
| Excellent | 465/2021 | (23.0) | 27/267 | (10.1) | 96/725 | (13.2) | 36/314 | (11.5) |
|  | Freq | (%) | Freq | (%) | Freq | (%) | Freq | (%) |
| Very good | 813/2021 | (40.2) | 84/267 | (31.5) | 247/725 | (34.1) | 115/314 | (36.6) |
| Good | 570/2021 | (28.2) | 95/267 | (35.6) | 256/725 | (35.3) | 122/314 | (38.9) |
| Fair | 134/2021 | (6.6) | 49/267 | (18.4) | 108/725 | (14.9) | 36/314 | (11.5) |
| Poor | 39/2021 | (1.9) | 12/267 | (4.5) | 18/725 | (2.5) | 5/314 | (1.6) |
| **Life satisfaction: happy with life as a whole** |  |  |  |  |  |  |  |  |
| Happy | 1938/2007 | (96.6) | 238/263 | (90.5) | 668/709 | (94.2) | 298/311 | (95.8) |
| Unhappy | 69/2007 | (3.4) | 25/263 | (9.5) | 41/709 | (5.8) | 13/311 | (4.2) |
| **Life satisfaction: happy with the future** |  |  |  |  |  |  |  |  |
| Happy | 1923/1978 | (97.2) | 228/252 | (90.5) | 653/689 | (94.8) | 288/302 | (95.4) |
| Unhappy | 55/1978 | (2.8) | 24/252 | (9.5) | 36/689 | (5.2) | 14/302 | (4.6) |
| **Completion of high school (Year 12 or Cert II)** |  |  |  |  |  |  |  |  |
| Completed Year 12/Cert II | 1978/2028 | (97.5) | 254/268 | (94.8) | 706/731 | (96.6) | 296/316 | (93.7) |
| Did not complete | 50/2028 | (2.5) | 14/268 | (5.2) | 25/731 | (3.4) | 20/316 | (6.3) |
| **Completion of any post-school qualification** |  |  |  |  |  |  |  |  |
| Yes | 1741/2028 | (85.8) | 215/268 | (80.2) | 629/731 | (86.1) | 264/316 | (83.5) |
| No | 287/2028 | (14.2) | 53/268 | (19.8) | 102/731 | (13.9) | 52/316 | (16.5) |
| **Participation in the labour force** |  |  |  |  |  |  |  |  |
| Employed | 1872/1927 | (97.2) | 233/242 | (96.3) | 652/686 | (95.0) | 281/290 | (96.9) |
| Unemployed | 55/1927 | (2.8) | 9/242 | (3.7) | 34/686 | (5.0) | 9/290 | (3.1) |

In terms of educational attainment at age 25, membership in the *increasing exercisers* group was associated with lower odds of year 12 high school completion, compared with *regular weekly* exercisers (adjusted OR for *increasing exercisers* =0.43, 95% CI 0.23, 0.80; p<.008)(Fig 20). Young people in the *decreasing exerciser* group were less likely than *regular weekly* exercisers to participate in the workforce at age 25 (adjusted OR for *decreasing exercisers* =0.58, 95% CI 0.35, 0.95; p<.031) (Fig 22). There were no other associations of exercise trajectory membership with measures of educational or occupational attainment.

## 4. Discussion

To our knowledge, this study is the largest longitudinal investigation to date of long-term recreational exercise patterns in young people during the transition from adolescence to early adulthood. Using GBTM, we delineated four distinct patterns of exercise behaviours over the 8-year period from age 16 to 24, for each of two definitions of exercise engagement. Factors predicting long-term adherence to daily recreational exercise as recommended by WHO guidelines were male gender, higher levels of academic self-efficacy, higher levels of sports participation, less time spent watching TV, identifying as indigenous, and lower parental socioeconomic status (SES). Higher academic literacy at age 15 was associated with a decreased likelihood of adhering to WHO guidelines over the subsequent 8 years, and with increased odds of reducing exercise from daily levels, over time.

Conversely, factors at age 15 predicting highly irregular (less than *once weekly*) exercise over time were female gender, lower levels of academic self-efficacy, lower levels of sports participation, more time spent watching TV, and lower parental socioeconomic status (SES). Higher academic literacy at age 15 increased the risk of falling into both the *declining exerciser* and the *increasing exerciser* groups.

At age 25, participants in the *guideline* exercise trajectories were more likely to report better general health, compared with those who never met guideline frequency (model 1), while young people with persistent irregular exercise of less than once weekly frequency, were more likely to report poorer outcomes in measures of general health, mental health, and life satisfaction compared to the more regular (at least once weekly) exercisers (model 2) (Fig 3). Measures of educational attainment or participation in the labour force were not associated with exercise trajectory membership.

Earlier studies applying GBTM to primary school children to about 18 years of age found evidence that latent classes of exercise behaviours with best model fit were of 2 to 4 groups in size [33, 34]. Overall, these studies suggest that physical activity levels decline from childhood to the age of about 16 in most children. Our model 2 indicated that between the ages of 16 and 24, self-reported exercise patterns were relatively stable at a modest level (once weekly or more) for a majority of participants, consistent with other studies in older adolescents reporting that the majority clustered within stable trajectories of low-to moderate exercise frequency [35]. The notion that long-term exercise habits are already quite firmly established by late adolescence is further supported by studies that followed young adults into mid-adulthood, showing that most participants were either stably “moderately active” [36], “persistently low active” [37], or “inactive” [38]. Notably, the latter two studies also support our finding of discernible smaller groups who show changes in exercise behaviours in late adolescence and adulthood (i.e., “increasers” and “decreasers”).

Our finding that female gender is the strongest risk factor for lower or less consistent recreational physical activity between the ages of 16 and 24 (Exercise Trajectory Models 1 and 2) echoes previous reports that young females are generally less physically active than males [39], and that they are at higher risk of decreasing activity levels over time [40]. Multiple factors have been reported that put young females at disadvantage, including reduced opportunity, lower access, and lack of sports diversity, but also divergent parental and cultural expectations, stereotypes, and role models [41]. Psychological factors such as perceived sports competency may play an additional role [42]. Therefore, active encouragement and validation of female sports participation across institutions and media are required to counteract the dropout of female adolescents from regular recreational physical activity.

We identified that higher academic self-efficacy at age 15 predicted higher and more consistent long-term exercise behaviours. Self-efficacy is an individual’s belief in their ability to perform a specific action required to attain a desired outcome, and is thought to lead to setting higher goals, investing more effort in the pursuit of these goals, and persisting through barriers or setbacks [43]. In adolescents and young adults, associations between high levels of self-efficacy and higher physical activity levels have been shown [44], and self-efficacy is thought to be one of the driving cognitive factors for engagement in regular exercise [45]. Psychological health interventions focusing specifically on self-efficacy have shown some promise in improving exercise engagement in this age group in the short term [46]. Our findings suggest that such interventions, delivered around the age of 15, may also have a positive impact on the establishment of long-term exercise behaviours.

Time spent exercising and time spent watching television at age 15 were additional predictors for participation in physical exercise. Previous studies have found that sports participation in adolescence predicts physical activity in adulthood [15, 47]. An inverse relationship between longitudinal patterns of television viewing and physical exercise in teenagers has also been reported previously [48]. Our findings underscore the importance of establishing regular recreational exercise habits in early adolescence, and of limiting screen-based leisure activities, to prevent ongoing inactivity or decline.

While parental socioeconomic factors have been reported as important for exercise engagement in children and younger adolescents [34], we found they had less impact in our older, more independent cohort of 16-24 year-olds. Our finding of higher academic literacy as a risk factor for lower and less consistent longitudinal exercise groups is interesting, and perhaps reflects competing academic demands (e.g., university entry). Increased exercise opportunities for high academic achievers, for example through programmes in senior high school and university, should be considered.

The favourable outcomes at age 25 for young people engaging in at least once weekly recreational exercisers are in keeping with previous prospective studies in adult populations that have shown benefit from exercise in measures of physical [49] and mental health [9]. In adolescents and young adults, previous longitudinal studies have found advantages for regular or increasing exercisers for educational outcomes [50] and for health-related quality of life measures [14]. Recently, the first longitudinal study reporting objectively measured physical activity in 12-18 year-olds convincingly demonstrated protective effects against depressive symptoms at age 18 [35]. In contrast, one study reported that frequent physical exercise at age 14 was not associated with affective, anxiety, or substance use disorder diagnoses at age 21[51]. We argue that trajectory analysis of repeatedly-measured data provides a more accurate measure of behaviour patterns over time and is hence more sensitive for predicting longitudinal outcomes. As might be expected, outcome differences between those reporting ongoing daily exercise (“guildeline”) and those with less regular regimes were less pronounced. Nevertheless, long-term daily exercisers reported significantly better subjective general health at age 25, an age where physical health concerns are not particularly prevalent. We would expect that these subtle health advantages of daily exercise increase in older age groups.

### 4.1 Strengths and Limitations

We used data from a national prospective cohort study with a large sample size and follow up over 8 years. As expected, participation rates declined over the course of the study, and non-random dropout may be a source of bias. Supplementary Table 1 provides a comparison of baseline characteristics and trajectory group membership between those with missing outcome data and those with outcome data available. Exercise data were self-reported and are likely less reliable than more objective measures of exercise participation such as wearable actigraphy devices [52]. Objective monitoring of exercise over long periods such as the 8 years covered in our study will become more feasible in the future as wearable technology becomes increasingly widespread. The design of the exercise questionnaire used for LSAY led to skewing of categories towards very low exercise frequencies and prompted us to dichotomize exercise behaviours into binary variables used in two separate longitudinal models. While we believe that this approach captures behaviours most relevant from a public health perspective (adhering to international guidelines – model 1; and very irregular exercise engagement - model 2), future iterations of population studies such as LSAY should consider more precise scales to measure frequency of recreational exercise. We are unable to rule out residual confounding by factors not assessed in the survey with respect to the predictors of trajectory-group assignment and/or associations with the selected outcomes. These may include motivational, psychological or physiological resilience factors or life events that predispose individuals to particular patterns of exercise and achievement over the lifespan.

Our study design does not allow for conclusions about a causal relationship between exercise and our reported outcomes. It is possible that young people who enjoy better health, mental health, and educational success are also more prone to regular recreational exercise, or that more regular exercise is associated with other health behaviours that moderate our reported effects, such as smoking [53].

### 4.2 Implications of findings

Our findings could help inform preventive health initiatives in late adolescence and early adulthood and are applicable to high-school-, university-, and vocational college settings. While a higher level of consistent recreational exercise was associated with superior long-term outcomes across the measured domains, our findings also indicate that young people who increase exercise participation over time to even moderate levels (i.e., once weekly or more) experience advantages compared to those who are consistently inactive or whose participation decreases over time. Hence, the commencement and maintenance of regular exercise should be recommended and promoted at all ages. Our findings point to subgroups of young people who might benefit from more tailored exercise-promotion interventions. These include females, those with lower levels of self-efficacy, those who are inactive at age 15 with high screen time, those from lower SES families, and high academic achievers. Future research needs to identify the specific barriers and enablers of exercise participation for these groups.

## 5. Conclusions

Patterns of recreational exercise participation in late adolescence and early adulthood can be modelled as distinct latent trajectories. High school, university, and vocational colleges are important settings to implement interventions that support young people in establishing regular physical activity. Our study identified subgroups of young people that may especially benefit from targeted interventions. We showed that regular long-term regular exercise participation during the transition to adulthood may be associated with measurable benefits for health and mental health in young adults.

## Data Availability

The data underlying the results presented in the study are publicly available after an online registration and application process (www.ada.edu.au/).

## Acknowledgements

The authors would like to acknowledge the contribution of Tasman Swanton, Cameron Forrest, Emerick Chew, and Craig Fowler at NCVER.

## Declarations

The authors have no competing interests to disclose.

